# Language models reveal evidence gaps in variants of uncertain significance

**DOI:** 10.64898/2026.02.28.26347206

**Authors:** Weijiang Li, Vineel Bhat, Tian Yu, Matthew Lebo, Marinka Zitnik, Christopher Cassa

## Abstract

**Background:** Most rare coding variants in monogenic disease genes remain classified as Variants of Uncertain Significance (VUS), limiting their use in clinical care. Many variant classifications have been submitted to ClinVar, often with rich free-text summaries of the evidence underlying each classification. These narratives are not standardized and are difficult to mine systematically, making it challenging to identify variants that might be reclassified as new evidence becomes available.

**Methods:** We developed a two-stage language-model pipeline that (i) detects whether functional, population, or computational evidence is described in ClinVar and ClinGen variant summaries, and (ii) classifies whether it is evidence of pathogenicity or benignity. We first constructed Variant Evidence Text Annotations (VETA), a dataset of 44,522 ACMG/AMP keyword-description pairs derived from 18,678 ClinVar and ClinGen variant summaries using an LLM-based consensus annotation procedure. We then fine-tuned BioBERT-large models for each evidence type and stage, and validated performance using independent ClinGen expert-curated summaries as well as orthogonal variant-level evidence, including functional screening, computational scores, and population estimates of disease impact.

**Results:** Across evidence types, our models accurately identify whether functional, population, and computational evidence is present and whether it leans toward a pathogenic or benign impact. We find high agreement with ClinGen expert annotations and highly significant separation of validation scores between model-predicted benign and pathogenic groups (functional assays ***p* = 8.13 *×* 10*^−^*^30^**, variant allele frequencies ***p* = 4.11 *×* 10*^−^*^22^**, computational predictions ***p <* 8.88 *×* 10*^−^*^16^**). We applied the full workflow to approximately 6,000 ClinVar VUS variants whose submission summaries lacked explicit functional or population evidence. By aggregating external functional, population, computational, and diagnostic evidence using the ACMG/AMP SVI point-based framework, we found that about 17% of these VUS meet quantitative thresholds for a likely benign or likely pathogenic classification, including 492 VUS in genes reviewed by ClinGen Variant Curation Expert Panels.

**Conclusions:** Transforming unstructured variant summaries into a structured, evidence-type matrix enables scalable detection of evidence gaps, allowing for the systematic integration of new data sources, and prioritization of VUS that are most likely to be reclassified. This language model-enabled pipeline provides a generalizable digital approach to identify clinical evidence gaps as functional screens, biobank resources, and computational predictors continue to evolve.

## Introduction

High-throughput genomic sequencing has rapidly broadened our understanding of the phenotypic impact of human genetic variation. Yet a critical bottleneck remains in clinical care: many rare coding variants still lack sufficient evidence to be classified as pathogenic or benign with confidence. From 2019 to 2024, clinical laboratories deposited more than 3.6 million classifications in ClinVar, a public archive of genotype–phenotype relationships,[1], but many missense variants in clinically actionable genes remain labeled as Variants of Uncertain Significance (VUS) [2]. Because VUS do not provide clear guidance for management, they are generally not returned to patients outside of specific diagnostic contexts[3].

Variant interpretation is standardized through the American College of Medical Genetics and Genomics/Association for Molecular Pathology (ACMG/AMP) Sequence Variant Interpretation (SVI) framework, which defines multiple lines of evidence for determining whether a variant is pathogenic or benign[4]. These evidence types include population frequency (for example, PM2 for rare variants; BA1/BS1 for higher allele frequencies), case-level observations (PS4, BS2), functional assays (PS3, BS3), and computational predictions (PP3, BP4), among others. Expert curators weigh the strength of each evidence type, generally within a quantitative or Bayesian framework, to arrive at a final classification [5].

Over the last decade, more than two million ClinVar classifications have been accompanied by free-text variant summaries[6]. These narratives often include detailed ACMG-style evidence, such as functional screening evidence or case counts, but they are heterogeneous and rarely linked to structured evidence codes. As a result, laboratories and expert panels cannot systematically query the types of evidence considered or identified for each variant. They also cannot easily identify where one new source of evidence, such as a functional assay or updated computational score, might change a classification. Without this structured information, new evidence may be underutilized, because expert re-interpretation is resource intensive and does not scale with the volume of new data.

ClinGen has been addressing this challenge through Variant Curation Expert Panels (VCEPs), disease- and gene-focused groups that adapt ACMG/AMP guidance and provide standardized expert interpretations to improve consistency across laboratories. Because VCEP review is inherently expert-driven, panels cannot curate all variants and instead use explicit prioritization workflows to focus effort where it is most likely to impact clinical care. For example, ClinGen prioritizes curation of (i) medically actionable interpretation conflicts in ClinVar, (ii) variants repeatedly submitted as VUS by multiple laboratories, (iii) variants with a strong likelihood of definitive reclassification based on clear mechanistic or frequency evidence, and (iv) nominated variants of particular clinical or research importance[7]. In this context, scalable methods that identify variants with missing or newly available evidence can complement established prioritization criteria by helping expert panels and laboratories target variant review to those most likely to move out of the VUS category.

Recent work has explored the use of large language models to improve variant interpretation, including approaches that extract related evidence from the literature [8] or augment LLMs with curated databases to generate more accurate variant summaries [9]. In this work, we introduce a language model-enabled pipeline that converts free-text variant summaries into a structured matrix of ACMG/AMP evidence types.

We developed a two-stage framework that (i) detects whether functional, population, or computational evidence is described in a classification text summary and (ii) classifies whether each evidence type indicates a pathogenic or benign impact. We then use these classifiers to identify VUS whose summaries lack functional or population evidence, and analyze them using newly available data, including updated functional assays in MaveDB [10] or disease enrichment at the variant level within the UK Biobank [11]. This approach reveals thousands of VUS that are currently likely missing important lines of evidence in their text summaries. Importantly, our goal is not to automate variant classification, but to enable scalable detection of missing or newly available evidence so that expert review can be focused where reclassification is most likely.

## Results

### Developing a training dataset of evidence annotations

We first constructed Variant Evidence Text Annotations (VETA), a large corpus of ACMG evidence annotations derived from variant summaries within ClinVar and ClinGen (Figure 1a). From ClinVar, we identified submission-level summaries containing explicit ACMG/AMP evidence codes and used an LLM-based annotation pipeline to extract the corresponding descriptive text for each code. From ClinGen, we leveraged expert-curated variant summaries from the ClinGen Evidence Repository ^1^, where ACMG codes and supporting text are already linked. Across both sources, this procedure yielded 82,311 initial keyword-description pairs.

**Fig. 1:**
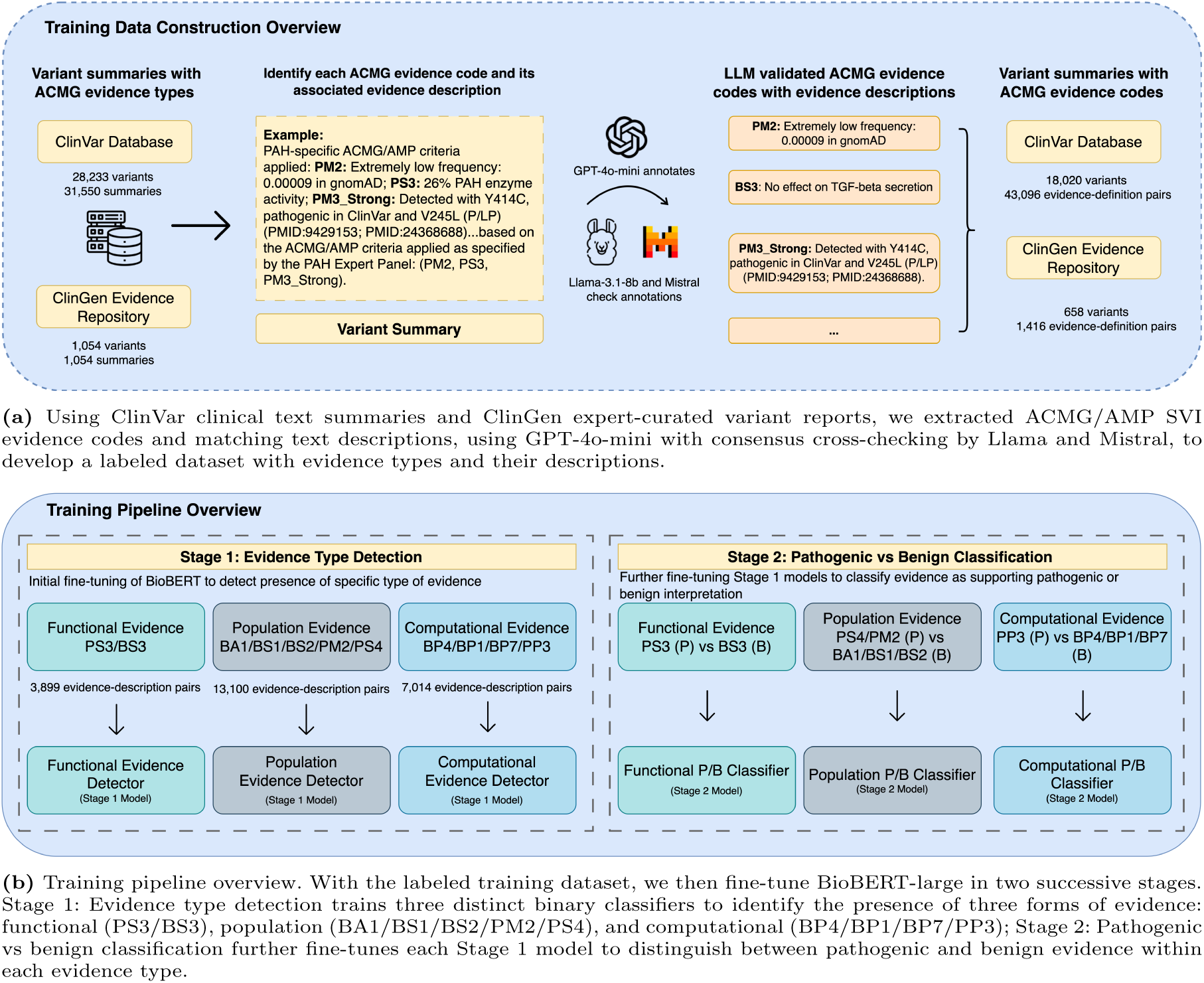
Overview for constructing a labeled dataset for ACMG evidence, and overview of the training pipeline. **(a)** Using ClinVar clinical text summaries and ClinGen expert-curated variant reports, we extracted ACMG/AMP SVI evidence codes and matching text descriptions, using GPT-4o-mini with consensus cross-checking by Llama and Mistral, to develop a labeled dataset with evidence types and their descriptions. **(b)** Training pipeline overview. With the labeled training dataset, we then fine-tune BioBERT-large in two successive stages. Stage 1: Evidence type detection trains three distinct binary classifiers to identify the presence of three forms of evidence: functional (PS3/BS3), population (BA1/BS1/BS2/PM2/PS4), and computational (BP4/BP1/BP7/PP3); Stage 2: Pathogenic vs benign classification further fine-tunes each Stage 1 model to distinguish between pathogenic and benign evidence within each evidence type.

To improve annotation quality, we applied a consensus filter using two independent instruction-tuned LLMs (Mistral-7b[12] and Llama-3.1-8b[13]) that evaluated whether each extracted text segment correctly matched the ACMG evidence code. We retained only keyword-description pairs that were confirmed by both models, resulting in 44,522 high-confidence annotations across 18,678 variant summaries that span all major ACMG evidence categories (VETA, Supplementary Table 2; model agreement rates, Supplementary Table 1).

With this annotated training dataset, we adopted a two-stage fine-tuning strategy built on BioBERT-large, as described in Figure 1b. Stage 1 focuses on evidence-type detection: we train three independent binary classifiers to recognize whether a narrative contains *functional* (PS3/BS3), *population* (BA1/BS1/BS2/PM2/PS4), or *computational* (BP4/BP1/BP7/PP3) evidence. Stage 2 predicts whether evidence detected in stage 1 is pathogenic vs. benign, with the goal of distinguishing pathogenic cues (e.g., PS3) from benign cues (e.g., BS3) within the same evidence type. We do this by further fine-tuning each stage 1 model using text samples that are labeled as containing each evidence type. This decoupled design leverages shared vocabulary within each evidence class while reducing label imbalance, and ultimately yields six classifiers whose output can be used to identify variants with these forms of ACMG/AMP evidence for each variant summary. Model performance results evaluated with the test split data after training are included in Supplementary Table 3.

### Evaluating model performance using expert-curated clinical summaries

We first evaluated the consistency of model predictions within VETA, using gene-stratified cross-validation, both Stage 1 (evidence type presence) and Stage 2 (evidence pathogenicity). All models achieved high accuracy and F1 scores for functional, population, and computational evidence (Supplementary Tables 3). Next, we evaluated models trained using only VETA annotations from ClinVar, and made model predictions on separately expert-curated ClinGen summaries. Using the complete ClinGen summary text, we again evaluated each model’s performance, checking whether each evidence type was present (Stage 1) and had the correct directional prediction (Stage 2) within each ClinGen summary. Models trained on ClinVar annotations generalized well to expert-curated ClinGen summaries, correctly identifying the presence of each evidence type and distinguishing pathogenic from benign evidence directions despite differences in how variant reports are structured within the two resources (Supplementary Table 4, Supplementary Tables 5-6). For population evidence, we additionally report results using ‘presence-verified’ labels, which account for evidence codes discussed in the text but which may not have met evidence standards for specific VCEPs (See Methods).

### Evaluating model performance using computational, functional, and population data

To further evaluate model validity, we analyzed Stage 2 predictions against independent quantitative benchmarks, including functional screening scores from FUSE [14], variant-level odds ratios of disease from the UK Biobank[11], gnomAD allele frequencies [15], as well as AlphaMissense[16] and REVEL [17] computational scores. For each variant text summary, we calculated model predictions for each evidence type. We grouped these prediction scores into the top quartile for summaries most likely to contain pathogenic evidence of that type, the bottom quartile for summaries most likely to contain benign evidence, and the middle two quartiles for summaries that are most uncertain. We then compared these model predictions against the corresponding externally-derived validation scores; Supplementary Figure 1 describes the numbers of genes and variants that have each form of evidence in each stage, for each evidence type.

We find a clear separation between model-predicted scores for each form of evidence, including computational scores (AlphaMissense), functional screening scores (FUSE), and allele frequency data (gnomAD) (Figure 2). All stage 2 models had robust classification performance when distinguishing pathogenic from benign evidence, across all three evidence categories (Supplementary Table 7), validating their ability to predict the direction of evidence from clinical text. In each case, model-predicted pathogenic evidence was associated with significantly more damaging quantitative scores than model-predicted benign evidence, supporting the ability of this text-based approach to make predictions consistent with biologically meaningful signals.

**Fig. 2:**
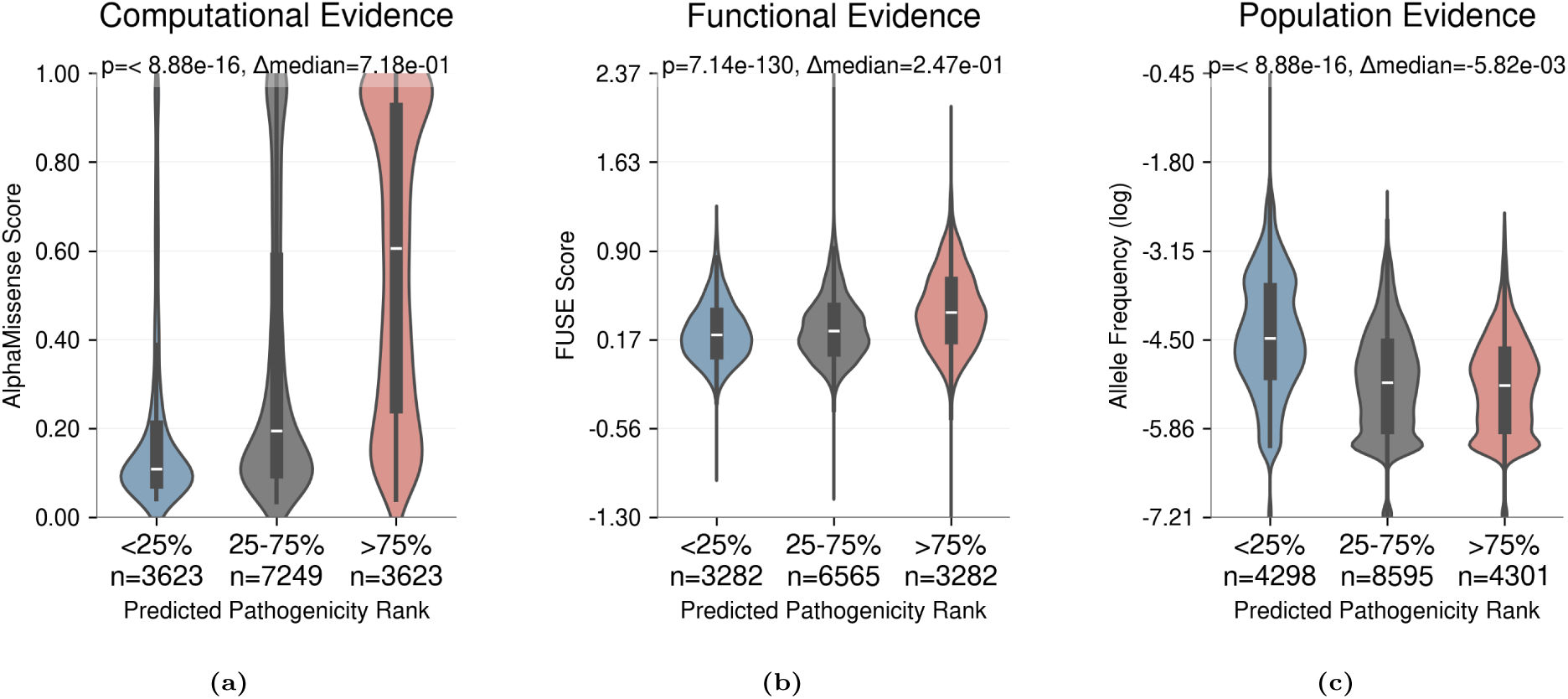
Aggregated Stage 2 model predictions stratified by percentile-based thresholds. *x*-axis: predictions assigned to pathogenic (top 25%), uncertain (middle 50%), and benign (bottom 25%); *y*-axis: model scores. (**a**) Computational evidence (AlphaMissense). (**b**) Functional evidence (FUSE). (**c**) Population evidence (allele frequency).

We find that these models can predict well across individual genes (Figures 3). For each form of evidence, we analyze the top 5 genes by variant count, as some genes do not have functional scores, and score availability can vary by variant type. Figure 3a shows the distribution of AlphaMissense scores for each variant summary based on its existing ClinVar submission classification label. We then make model predictions for all ClinVar VUS submissions, and plot our calibrated Stage 2 model predictions on the *x*-axis, and their corresponding AlphaMissense score on the *y*-axis (Figure 3b). Notably, the model-predicted distribution closely aligns with existing ClinVar classifications, with model-predicted categories showing comparable AlphaMissense score distributions to their corresponding ground-truth labels. Similar validation results for functional evidence are shown in Figures 3c, 3d.

**Fig. 3:**
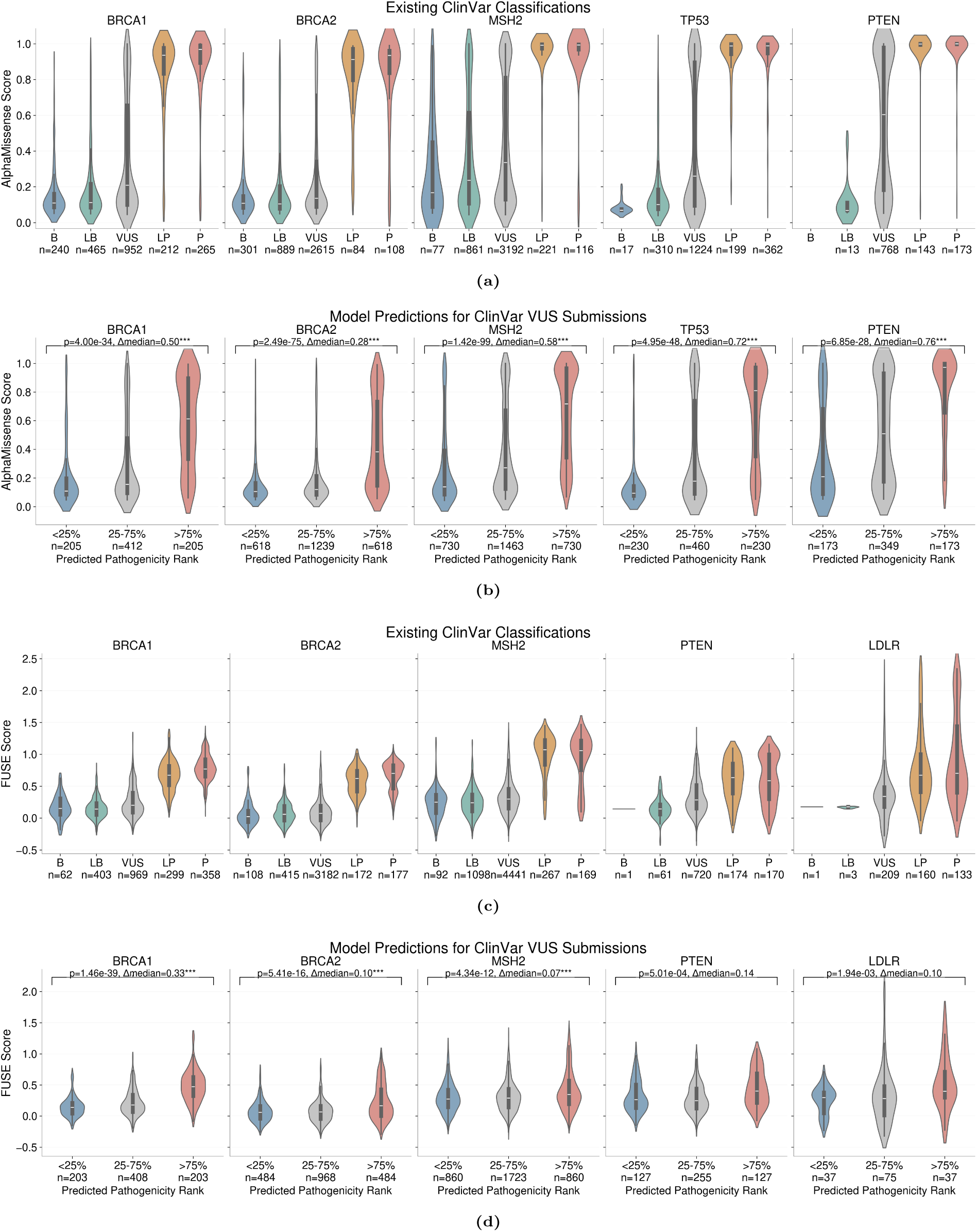
External validation of model predictions using orthogonal evidence scores. (**a, b**) Computational evidence: AlphaMissense scores across ClinVar classifications (a) and model-predicted VUS categories (b). (**c, d**) Functional evidence: FUSE scores across ClinVar classifications (c) and model-predicted VUS categories (d). VUS submissions were stratified into three percentile bins, ranked by predicted pathogenicity: low (bottom 25%), intermediate (middle 50%), and high (top 25%). Statistical significance reported after Bonferroni correction (* *p<*0.05, ** *p<*0.01, *** *p<*0.001). See Supplementary Figure 2 for population evidence (gnomAD allele frequency) validation.

Additional validation results for computational scores, allele frequencies, and variant-level odds ratios of disease are provided in Supplementary Figures 3 and 4 in Supplementary Material. Additionally, we evaluated thresholding our model predictions based on expected proportions of P/B/VUS variants from external scores in Supplementary Figures 5, 6, and 7 in Results for Alternative Thresholds, for the top 5 genes by variant count in each figure.

**Fig. 4:**
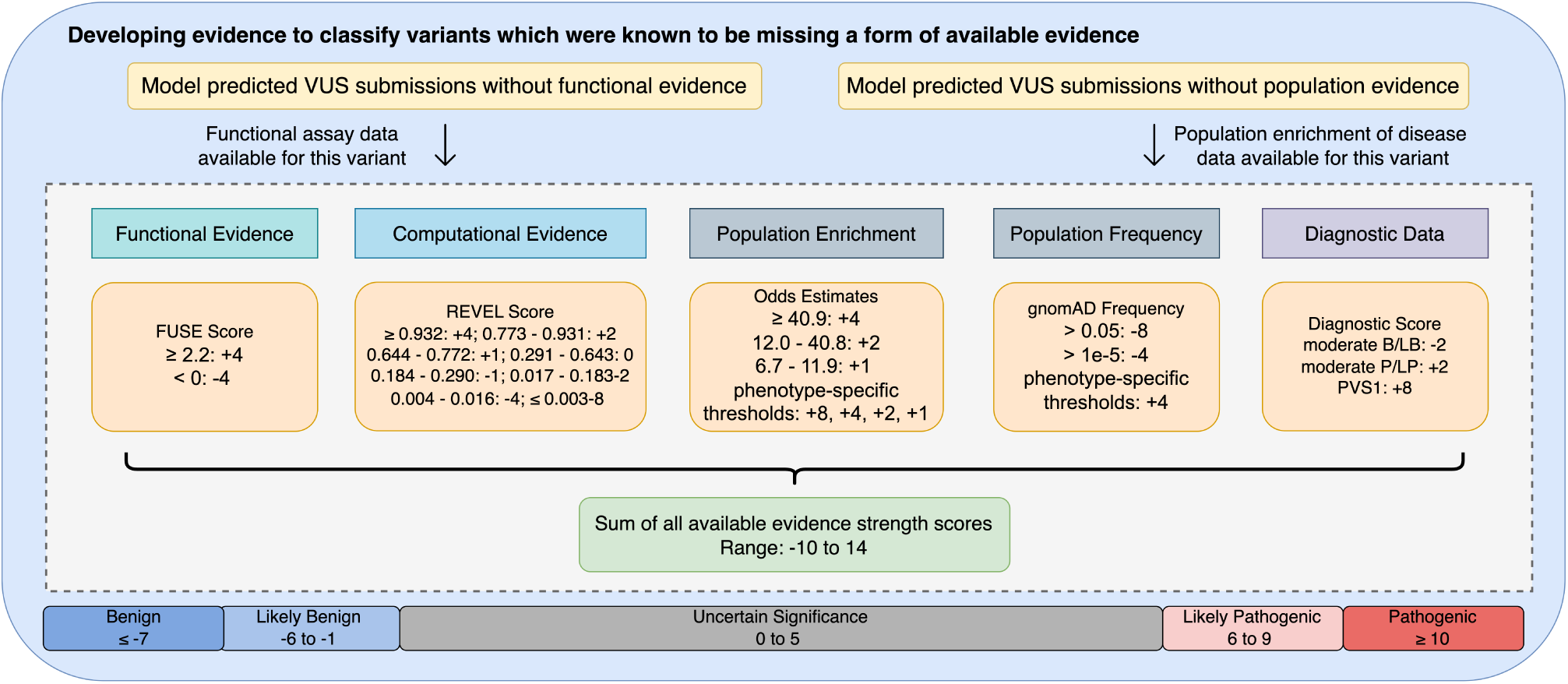
Overview of the external evidence scoring system for ClinVar VUS classification. The pipeline evaluates five categories of evidence: (1) functional evidence using FUSE scores; (2) computational evidence via REVEL scores; (3) population enrichment through odds estimates with phenotype-specific thresholds; (4) population frequency data using gnomAD frequencies with phenotype-specific thresholds; (5) diagnostic data incorporating diagnostic scores. Individual evidence scores are summed to produce a total evidence strength score ranging from -10 to +14, which maps to five variant classification categories: benign (*≤ −*7), likely benign (-6 to -1), uncertain significance (0 to 5), likely pathogenic (6 to 9), and pathogenic (*≥* 10). This quantitative approach leverages external evidence sources to classify ClinVar VUS variants.

### Model validation using an LLM external judge

Next, as an additional check of our model, we employed an LLM as an external judge of the consistency of our predictions with ACMG evidence type definitions, guided by prior work [18]. We observed high rates of agreement between our classifiers and Llama-3.1-8b, exceeding 90% in the highest-confidence prediction bins (confidence score *>* 0.9) across evidence types. Complete results across models are shown in Supplementary Figure 8. We further found that Llama-3.1-8b disagrees with our model predictions most frequently in cases where evidence descriptions were ambiguous. For example, our model would mistakenly consider the summary to contain a type of evidence when the text mentions the evidence type that was evaluated, but does not provide any additional specific details describing that evidence.

### Identifying evidence gaps in clinical summaries

Our next goal was to identify variants that are currently unclassified within ClinVar (VUS), where evidence might not have been previously considered, but is now available. We first used our functional and population evidence models to identify variants that are predicted to not contain either form of evidence. As a variant might have more than one submission record in ClinVar, we applied a variant consistency check to identify variants where all ClinVar submission records were predicted not to contain either form of evidence, as information might have become available in later submissions (details included in Variant-Level Evidence Classification Consistency Analysis). This filters out variants with conflicting evidence classifications by our classifiers: 379 variants (2.6%) were filtered for population evidence, and 1,714 variants (11.8%) were filtered for functional evidence.

Next, we sought to identify whether there is sufficient evidence to classify these variants that are predicted not to have considered functional or population evidence by our classifiers. This set contained 6,084 ClinVar VUS overall and 3,865 VUS with linked UK Biobank phenotype data. We integrated external functional, population, and computational information for these variants using an ACMG-compatible point-based scoring system that combines FUSE functional scores, REVEL computational scores, variant-level odds ratios from the UK Biobank, gnomAD allele frequencies, and diagnostic evidence (PS1/PM5/PVS1), as described in Figure 4). We then calculated the total number of evidence points for each variant, and variant classification labels were applied consistently with the ACMG/AMP SVI framework.

The resulting evidence strength scores spanned the full range of ACMG categories, from benign to pathogenic (Figure 5). Variants with higher evidence scores were carried by a larger proportion of affected UK Biobank participants, and variants in the likely benign or benign range showed substantially lower disease prevalence, supporting the clinical relevance of the score distribution. Overall, 1,082 of 6,070 variants (17%) that lacked functional or population evidence in their ClinVar summaries met criteria for classification as likely benign, benign, likely pathogenic, or pathogenic classifications under our quantitative scheme, potentially affecting 6,245 individuals who carry one of these variants within the UK Biobank.

**Fig. 5:**
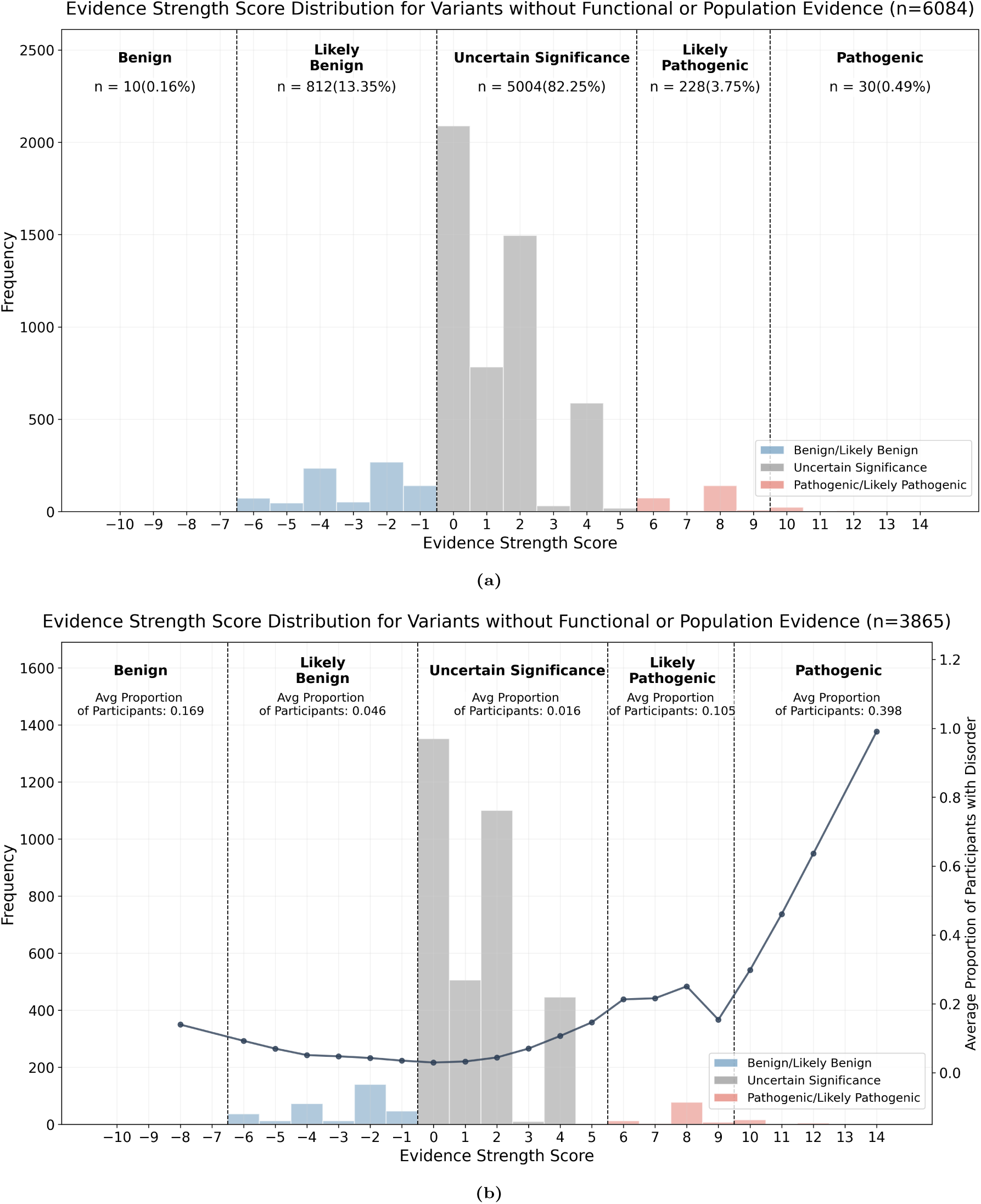
(**a**) Overall distribution of evidence strength scores across all 6,084 variants classified to have no functional or no population evidence by the functional evidence classifier and population evidence classifier. Variants are categorized into five clinical significance regions: benign, likely benign, uncertain significance, likely pathogenic, and pathogenic, based on a range of evidence strength scores. Vertical dashed lines indicate region boundaries. The histogram is colored by different clinical significance (P/LP, VUS, and B/LB). (**b**) Distribution of evidence strength scores across 3,865 variants classified to have no functional or population evidence by our classifier with phenotype data available. The curve is plotted with all data, with patient case data available, showing the proportion of participant correlation with evidence strength scores for variants without identified functional evidence (n = 3,865). The curve is smoothed by LOESS smoothing, which shows the average proportion of participants’ change across the histogram.

Outside of functional and population evidence, computational evidence was very often included in variant submission summaries, so evaluated it separately in Additional Results Figure 9a, and Figure 9b.

### Many variants within ClinGen Variant Curation Expert Panels have evidence gaps

ClinGen Variant Curation Expert Panels (VCEPs) apply gene- and disease-specific ACMG/AMP specifications and follow structured prioritization approaches to focus limited expert effort on variants where curation is most likely to have clinical impact (for example, variants with conflicting interpretations, variants repeatedly submitted as VUS, or variants likely to be definitively resolved with available data)[7].

To estimate the near-term opportunity for expert review within the VCEP-relevant scope, we intersected our “evidence-gap” VUS set with genes that are included on ClinGen expert curation panels. Among 2,347 ClinVar VUS in these genes, 492 (21%) met our quantitative thresholds for likely benign/benign or likely pathogenic/pathogenic classification after integrating external functional, population, computational, and diagnostic evidence. These variants represent a focused subset where expert review could plausibly yield immediate reclassification impact, and they provide a concrete starting point for VCEP workflows that must triage evaluation among many candidate VUS.

## Methods

### Developing a labeled set of ACMG evidence types and variant descriptions

Our goal is to build a reproducible digital pipeline that uses clinical text to identify ACMG/AMP evidence types and their directions across large variant repositories. We therefore first constructed a labeled dataset linking ACMG evidence codes (e.g., PM2, PS3, BA1) to the specific descriptive text used in ClinVar and ClinGen variant summaries.

Within ClinVar, we work from variant submission summaries that explicitly mention any of the evidence codes used in the 2015 SVI guidelines. From ClinGen, we make use of the expert-curated variant summaries in the ClinGen Evidence Repository (https://erepo.clinicalgenome.org/evrepo/). Within ClinVar, when we identified any specific SVI keyword, again, e.g., ‘PM2’, our goal is to find additional text that describes that specific form of evidence, e.g., *”this variant is observed to happen in 0.05% of the population”*. By working from a dataset where these evidence keywords are explicitly included, we could be more confident that there would be an association description of the review of that form of evidence. For the ClinGen Evidence Repository, it was more straightforward to extract these keyword-description pairs, as they are provided explicitly along with each variant summary. We then use all of these ClinVar and ClinGen samples as the corpus for developing a set of LLM-based keyword-description annotations.

To identify the associated text for each SVI evidence keyword, we used GPT-4o-mini [19] to identify potential text descriptions related to each specific evidence type. We provided GPT-4o-mini each summary along with three examples as the prompt (LLM Annotation Prompt). Then we used a consensus-based process using Llama-3.1-8b [13] and Mistral-7b [12] to check the annotation results from GPT-4o-mini, and only retained annotations that both Llama and Mistral agreed on. We used a temperature = 1.0 to query GPT-4o-mini for the initial annotation, and a temperature = 0.0 to query Llama and Mistral for checking annotation results, and the agreement rates are included in Supplementary Table 1. We named the resulting dataset **VETA** (Variant Evidence Text Annotations), an annotated dataset comprising 44,522 labeled examples derived from 18,678 variant summaries. Detailed information about the dataset is described in VETA Dataset Annotation and Distribution Statistics.

### Training Evidence Classifiers

With the labeled data, we then fine-tuned BioBERT-large [20], a language model pre-trained on a biomedicine domain-specific corpus. BioBERT is domain-specific pre-training on PubMed abstracts and PMC full-text articles, making it well-suited for biomedical text classification tasks.

#### Data Processing

We applied data preprocessing to improve data quality and model performance, including removing duplicate descriptions for the same ACMG code, removing text data that contains fewer than 10 words, removing references to publications, and removing non-English words. To ensure we have a diverse text corpus for training, we performed deduplication using MinHash [21] with a similarity threshold of 0.95. By doing so, we removed nearly duplicated data examples while maintaining diverse training data examples for the same ACMG codes.

For functional evidence classification, we addressed the challenge of implicit negation in clinical text through pattern-based negation detection. We designed regular expression patterns to identify statements that explicitly deny the existence of functional evidence, including phrases such as “no functional evidence” and “failed to demonstrate functional effect”. Text containing such negation patterns was classified as negative examples during Stage 1 training to improve model specificity.

Some evidence types, such as PM2, appear more frequently than others. Thus, we addressed the class imbalance issue by stratified sampling. For overrepresented classes, we applied random undersampling to achieve a class ratio of 2:1 pathogenic to benign ratio.

### Two-Stage Classification Framework

We implemented a two-stage inference approach to identify evidence presence and determine pathogenicity across three ACMG evidence categories: functional, population, and computational.

#### Model Architecture and Evidence Mappings

Stage 1 models function as binary classifiers distinguishing summaries containing specific evidence types from those without. Stage 2 models, built upon Stage 1 architecture, classify evidence direction with the following pathogenicity mappings:

- **Functional**: PS3 (pathogenic) versus BS3 (benign)
- **Population**: PS4/PM2 (pathogenic) versus BA1/BS1/BS2 (benign)
- **Computational**: PP3 (pathogenic) versus BP4/BP1/BP7 (benign), where BP7 specifically addresses synonymous variants with predicted splicing effects

#### Sentence-Level Inference Pipeline

For all evaluations and applications, we process clinical text at the sentence level using the following standardized pipeline:

##### Text Preprocessing

We apply our previously trained SentenceClassifier from ClinVar-BERT [6] to label sentences as description, evidence, or conclusion, retaining only evidence-labeled sentences for classification. Sentences shorter than 10 words are excluded from analysis.

##### Evidence Detection and Classification

Stage 1 models evaluate each evidence sentence independently. A variant summary is classified as containing evidence if any constituent sentence receives a confidence score *>* 0.5. For evidence-positive summaries, Stage 2 models determine pathogenicity, with the highest-confidence sentence prediction establishing the final classification (benign or pathogenic).

##### Summary-Level Aggregation

If at least one sentence contains specific evidence, the entire summary is labeled as containing that evidence type. Summaries with no evidence-positive sentences are classified as lacking that evidence type.

### Model Evaluation

We evaluated our two-stage classification framework through three approaches: (1) internal validation using gene-stratified cross-validation, (2) independent evaluation on expert-curated ClinGen data, and (3) external validation against quantitative evidence scores. All evaluations used the same inference method described above.

#### Internal Validation

##### Gene-Stratified Cross-Validation

To assess model generalization across genetic loci, we implemented 5-fold cross-validation with gene-level stratification. We partitioned all genes into five non-overlapping folds, training on four folds (80% of genes) and evaluating on the held-out fold (20%). This approach ensures the model’s ability to generalize to unseen genes rather than memorizing gene-specific patterns. Performance metrics were aggregated across all five folds to provide robust estimates of model performance.

#### Keeping Evidence-Only Text for Evaluation and Clinical Application

In our previous work, ClinVar-BERT [6], we trained a SentenceClassifier that labels sentences in ClinVar summaries into description, evidence, and conclusion. To ensure that our evidence classification models can make the most accurate prediction, we used the SentenceClassifier to label sentences in our evaluation data, and we only kept sentences that were labeled as ‘evidence’. This applies to all evaluation and downstream application tasks mentioned in the following sections.

#### Independent Evaluation with ClinGen Expert-Curated Data

To validate performance on high-quality expert annotations, we evaluated models trained exclusively on ClinVar VETA annotations using the ClinGen Evidence Repository as an independent test set. This cross-repository evaluation required careful handling of structural differences between data sources.

##### Data Extraction and Processing

We extracted clinical text from ClinGen’s ‘Summary of interpretation’ column and ground truth labels from the ‘Applied Evidence Codes (Met)’ field. We recognize that there are fundamental differences in summary structure between these datasets. ClinGen summaries are evidence-focused with explicit ACMG/AMP criteria listings (e.g., “PAH-specific ACMG/AMP criteria applied: BA1, BP5”), while ClinVar summaries emphasize descriptions and variant-level assertions. We filtered sentences containing “ACMG/AMP criteria applied” to prevent simple pattern matching and ensure genuine evidence detection from clinical descriptions.

To address potential false positive patterns while maintaining evaluation independence, we partitioned ClinGen data into 20% validation and 80% test sets using stratified sampling by evidence type. The validation set served exclusively for identifying systematic error patterns (particularly for handling negation, which our VETA dataset does not cover) to inform targeted synthetic training data generation, while the test set remained strictly held out for final evaluation.

##### Training Data Augmentation

Based on our analysis of the ClinGen validation data, we observed that negation is a primary source of false positives. We used the validation data as examples to prompt GPT-4o-mini to generate synthetic negation examples following observed patterns (e.g., “These studies do not meet the criteria set forth by the MDEP for the application of PS3 or BS3 (PMID: 21170474)”, “does not meet our threshold criteria for PM2 supporting or BS1”, etc.). These synthetic examples (n=250) were added to the VETA training data to improve negation handling without exposing the model to ClinGen test data.

##### Stage 1 Evidence Detection Evaluation

For Stage 1 evaluation on the held-out ClinGen test set, summaries were labeled positive for an evidence type if any corresponding ACMG codes were present in the ‘Applied Evidence Codes (Met)’ field. For example, PS3 or BS3 codes indicated functional evidence. Models predicted binary presence/absence for each evidence type, with performance measured against these expert-determined labels.

For population evidence, some ClinGen text summaries include evidence that has been evaluated (e.g., PM2) but that a VCEP did not include in the Applied Evidence Codes section due to specialized guidelines or rules related to PM2 evidence. Because Stage 1 evaluates the presence of evidence, these samples should be labeled positive when the evidence code is explicitly mentioned in the text. For the purposes of Stage 1 classification, when this evidence was described and considered, but did not meet application thresholds and appeared in the “Applied Evidence Codes (Not Met)” column, we treated these separately. We included these in a separate group called ‘presence-verified’ labels, and if there is an explicit mention in the summary text that this form of population evidence had been discussed, it is labeled as a positive for that group. We verified code presence using a regular expression pattern matching for text matching ‘PM2’, including “Not Met” codes in the positive labels.

##### Stage 2 Evidence Direction Evaluation

Stage 2 evaluation used the same text processing but with directional labels derived from specific ACMG codes. We filtered summaries to include only those with relevant evidence codes, then assigned pathogenic or benign labels based on the specific codes (e.g., PS3: “pathogenic functional evidence”, BS3: “benign functional evidence”). This evaluated whether models trained on ClinVar’s assertion-focused text could accurately extract evidence direction from ClinGen’s evidence-focused summaries.

### External Validation Against Quantitative Evidence Scores

#### Validation Data Processing

To measure the performance of each evidence extraction model, we consider four complementary sources of external validation data. Below, we describe our sources of validation data for each evidence type:

##### Population-based evidence

We used population-based data: variant-level allele frequency data from gnomAD. Variant allele frequencies were used from the gnomAD v4.1 exome dataset, accessed via the gnomAD DB plugin

##### Functional evidence

We used FUSE variant functional impact scores for 232,104 missense variants spanning 48 genes, integrating deep mutational scanning data from MaveDB [10] and ProteinGym [22]

##### Computational evidence

We employed two in silico predictors: REVEL, an ensemble meta-predictor optimized for rare disease variants [17], and AlphaMissense, a deep-learning model trained on protein structural context and evolutionary conservation [16]. To mitigate potential data leakage, we validated against ClinVar submissions that were last updated before September 2023 (before AlphaMissense was published), when using AlphaMissense as the validation data.

#### Threshold-Derived Evidence Labels for Evaluation

Since explicit labels for evidence direction (pathogenic vs. benign) are rarely available in clinical submissions, we validated Stage 2 models using threshold-derived labels generated from quantitative evidence scores. These labels serve as proxy ground truth by applying clinically-established thresholds to convert continuous scores into categorical evidence directions:

- **Computational evidence**: REVEL *≥* 0.644: pathogenic; REVEL *≤* 0.290: benign
- **Functional evidence**: FUSE *≥* 1*σ*: pathogenic; FUSE *≤* 0: benign
- **Population evidence**: Odds ratio *>* 5 with CI *>* 1: pathogenic; Odds ratio *<* 1: benign

These threshold-derived labels provided ground truth for evaluating Stage 2 predictions. To address potential inconsistencies between threshold-derived labels and clinical classifications from submitting laboratories, we restricted our evaluation to ClinVar records where the evidence direction aligned with each testing lab’s classification result submitted to ClinVar.

#### Percentile-Based Confidence Thresholding

To select high-confidence predictions while maintaining gene-specific score distributions, we applied percentile-based thresholds within each gene. Percentile-based thresholds allow the use of gene-specific score distributions and avoid imposing a single absolute cutoff across genes with heterogeneous evidence availability. We ranked variants by their normalized evidence scores: 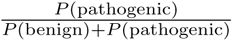 for pathogenic evidence and 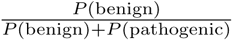 for benign evidence. Using a 25%/50%/25% split, we classified the top quartile as pathogenic evidence (P or PS3), the bottom quartile as benign evidence (B or BS3), and the interquartile range as uncertain. This approach ensures balanced representation across genes regardless of their baseline model confidence.

#### Statistical Analysis and Visualization

Model validation assessed the correlation between predicted evidence categories and quantitative evidence scores using statistical tests, and this validation is done at the submission level. We employed the Mann-Whitney U test to compare score distributions between opposing evidence categories: Likely vs Unlikely for Stage 1; PS3 vs BS3 or P vs B for Stage 2. In the visualization showing 5 genes, the p-value was adjusted by Bonferroni correction.

The full validation workflow and data distribution during the prediction process for each evidence model are illustrated in Figure 1. We only include ClinVar VUS submissions for the validation analysis. For population evidence, we only included rare variants with gnomAD allele frequency *≤* 0.5%, with consequences including missense, nonsense, or splice-affecting variants.

#### LLM Validation

To further validate model predictions, we leveraged LLMs to evaluate the accuracy of model predictions using the ACMG guideline [4] definition as the reference. This analysis is done at the variant submission level, where an LLM checks our model prediction of a variant submission record.

The prompt used for evaluation is included in LLM Validation Prompt. For each model prediction, we constructed evaluation prompts that provided both the ACMG evidence definition for the predicted evidence type and the specific variant summary from which the model made the prediction. LLMs were then asked to determine whether the extracted evidence was consistent with the provided ACMG definition, responding with “Correct” or “Incorrect” classifications. After running model predictions on the validation data through the LLM, we measure the agreement rate, which is the percentage of predictions from our trained model that have a ‘Yes’ response from the LLM, and the disagreement rate, which is the percentage of ‘No’ answers. We then made histograms of agreement rates and disagreement rates across all model confidence scores.

### Identifying Variants with Evidence Gaps

Furthermore, we aim to apply our trained evidence classifier for variant interpretation; thus, we developed a comprehensive framework that integrates multiple external sources of evidence on VUS variants that are identified by our model to not contain functional evidence or population evidence in the text summary.

For the purpose of this analysis, we identified VUS variants by restricting variants with ‘ClinicalSignificance’ being ‘Uncertain Significance’. Within these VUS variants, we used our evidence classification models to obtain variants that do not contain population evidence or functional evidence in the text summary.

#### Variant-Level Evidence Classification Consistency Analysis

Our classification models make predictions at the submission level, but downstream analyses require variant-level classifications. Since each variant may have multiple ClinVar submissions with potentially conflicting evidence predictions, we implemented an aggregation strategy to derive consistent variant-level classifications.

We applied the following rules to aggregate submission-level predictions:

1. **No evidence:** If all submissions for a variant were predicted to contain no evidence of a specific type, the variant was classified as having no evidence.
2. **Consistent evidence:** If one or more submissions were predicted to contain evidence, we examined whether all evidence-containing submissions agreed on the evidence type. Variants were considered consistent if all submissions with evidence were assigned the same category (e.g., all predicted as PS3, or all predicted as BS3, but no mixture of PS3 and BS3 for functional evidence). For consistent variants, we assigned the consensus evidence type as the variant-level prediction.
3. **Conflicting evidence:** Variants with contradictory evidence predictions across submissions (e.g., some submissions predicted as pathogenic and others as benign for the same evidence type) were excluded from downstream analysis to ensure reliability.

This aggregation approach prioritizes classification consistency and filters out ambiguous cases where submissions provide inconclusive evidence for the same variant.

#### Calculation of Evidence Score

We implemented a point-based scoring system following ACMG/AMP guidelines as demonstrated in Figure 4, where evidence strength corresponds to specific points: very strong (8 points), strong (4 points), moderate (2 points), and supporting (1 point) for pathogenic evidence, with corresponding negative values for benign evidence. This scoring framework allows the aggregation of multiple types of evidence.

##### Functional Evidence

Functional evidence uses FUSE scores derived from experimental assays. We set the thresholds for strong pathogenic as plus one sigma (standard deviation of FUSE score); variants with a FUSE score exceeding this threshold are assigned 4 points, while negative scores get -4 points as strong benign evidence. **Population Evidence** We incorporated two types of population evidence for variant classification. First, we used variant-level odds ratios quantifying disease enrichment for 5,737 rare (minor allele frequency *≤* 0.1%) coding variants across 41 clinically actionable genes covering 18 monogenic phenotypes, derived from UK Biobank clinical data [11]. These odds estimates provide phenotype-specific population context, with prevalence-adjusted thresholds for each phenotype detailed in Supplementary Material Table 8. Second, we analyzed allele frequency data from the gnomAD v4.1 exome dataset, accessed via the gnomAD DB plugin [15]. We applied phenotype-specific frequency thresholds listed in Supplementary Material Table 9 to determine variant rarity relative to expected disease prevalence.

##### Computational Evidence

For computational evidence, we used REVEL scores and applied strength thresholds that were developed in prior work [23]. These REVEL score thresholds describe the strength of evidence of pathogenicity or benignity for each possible REVEL score, ranging from supporting to very strong. Some REVEL scores do not provide evidence of pathogenicity or benignity. Full score thresholds and strength levels are provided in Supplementary Material Table 10.

##### Prior diagnostic evidence

We applied the Tocayo computational pipeline [24] to identify single-nucleotide variants that have structured evidence of pathogenicity based on existing variant classifications using the ACMG/AMP criteria PS1, PM5, and PVS1. This diagnostic evidence is based on variants with the same amino acid mutation. We integrated estimated moderate pathogenic diagnostic scores (+2 points), estimated moderate benign diagnostic scores (-2 points), and very strong pathogenic evidence (+8 points).

### Distribution of Evidence Strength Scores Across No-Evidence VUS Variants

To characterize the distribution of VUS variants across clinical significance categories, we analyzed the evidence strength scores for variants predicted to lack population or functional evidence by our classifiers. We applied established ACMG/AMP score thresholds [25] to categorize variants: scores *≤ −*6 as benign, *−*6 to *−*1 as likely benign, 0 to 5 as VUS, 6 to 9 as likely pathogenic, and *≥* 10 as pathogenic.

To visualize the relationship between the evidence strength score and the proportion of individuals with a positive phenotype case across different classification results, we employed LOESS (Locally Estimated Scatterplot Smoothing) regression analysis. LOESS is a non-parametric statistical method that fits smooth curves through data points without making assumptions about the global shape of the relationship. This approach uses locally weighted regression, where each bin’s average value contributes to the local fit based on its distance from the prediction point, with closer points receiving higher weights.

## Discussion

In this study, we developed an automated approach to extract and classify ACMG/AMP evidence from clinical variant text summaries. This addresses a bottleneck in identifying which forms of evidence have been included in variant interpretations, as many lab submission summaries do not enumerate the evidence keywords applied based on the ACMG/AMP SVI guidelines. By creating VETA, a large dataset of ACMG evidence annotations derived from ClinVar and ClinGen, and training BioBERT-based classifiers, we demonstrate that language models can identify functional, population, and computational evidence from free-form clinical text and determine whether that evidence leans toward a pathogenic or benign impact.

Our two-stage classification framework achieves strong performance across multiple validation approaches. Internal cross-validation on VETA demonstrated high accuracy and F1 scores for all three evidence types, and independent evaluation on expert-curated ClinGen summaries showed that models trained on ClinVar text generalize to structurally different narratives. External validation against orthogonal quantitative evidence scores further showed that text-derived evidence correlates closely with biological and clinical measures of variant impact, including functional scores, population-level odds ratios, allele frequencies, and computational scores.

We then embedded these classifiers into a broader pipeline that integrates external evidence in an ACMG-compatible point-based scoring framework to identify VUS with evidence gaps and reclassification potential. Among the 6,084 VUS variants our models classified as lacking explicit functional or population evidence in their ClinVar summaries (6,070 with complete scoring data), 1,082 (17%) met quantitative thresholds for benign, likely benign, likely pathogenic, or pathogenic classification when functional, population, computational, and diagnostic evidence were integrated. These variants represent high-priority candidates for expert review and potential reclassification. Notably, among 2,347 VUS within genes included on ClinGen Variant Curation Expert Panels, 492 (21%) reached at least likely benign or likely pathogenic thresholds, suggesting that focused curation efforts on these genes could yield immediate clinical impact.

New evidence sources will continue to emerge, including updated functional scores from multiplexed assays, expanded population evidence from biobanks, and improved computational predictors. Our evidence extraction framework enables systematic identification of variants that have not yet incorporated these lines of evidence in their clinical summaries. Rather than relying on manual review of thousands of records, which is not feasible at scale and is complicated by heterogeneous reporting styles, experts and clinical laboratories can use these model predictions to identify variant records missing specific evidence types and prioritize those variants for an updated review when new data becomes available.

As an example of the evidence gap in one gene, within *LDLR*, our model identified that 124 VUS lack functional evidence in their evidence text summaries. Among them, 122 potentially have newly available functional evidence from two recent functional assays [26, 27]. When considering this new evidence alongside other forms of available evidence, 19 can potentially be classified as B/LB and 4 as P/LP, when following the quantitative SVI framework (described in Identifying evidence gaps in clinical summaries.) For population evidence, only 39 *LDLR* VUS variants do not have population evidence mentioned in the text summary, and 3 potentially have new population evidence of pathogenicity in the form of an enrichment of elevated LDL cholesterol [11], and 2 can be classified as B/LB. These variants can be prioritized for review in descending order of probability that the relevant form of evidence exists from our models.

### Limitations of this work

This study has several limitations. First, our classifiers operate purely on text and therefore depend on the quality, completeness, and consistency of variant summaries. Summaries that mention that an evidence type was considered without providing specific details (e.g., “population evidence was reviewed”) can challenge these models. By using a two stage model, we have made headway at addressing this challenge. There are also many submissions that include some form of population evidence in the form of allele frequency or a computational score. Future models could be extended to identify specific outdated sources of evidence, such as small population datasets, where much larger datasets are now available, or outdated computational scores.

Second, our evidence extraction models make predictions at the submission level, and each prediction reflects the evidence described in a specific record. The prediction result only reflects the textual information for that submission, and the model does not automatically reconcile discrepancies across submissions or incorporate laboratory-specific practices, although we partially address this by reviewing consistency across submissions when we aggregate at the variant-level.

Third, there is some unavoidable circularity between the data used for training and validation. Because computational predictors and population frequencies are commonly used as evidence by clinical laboratories, similar information may be present in both the text summaries and the external resources used for validation. To mitigate this potential data leakage, we validated with ClinVar submission data that were last updated before AlphaMissense was published (September 2023) within the AlphaMissense validation plots, and used updated allele frequencies from gnomAD. However, given that many computational tools share related training sources, and that many sources of population frequency data are related, some data leakage within this validation remains possible. However, the overall strong agreement between our text-based predictions and sequence-based scores demonstrates that our approach effectively captures functional signals from clinical narratives.

Moreover, there can be mismatches between the specific types of evidence commonly used by testing laboratories and those available as new sources of evidence. For example, allele frequencies are the most commonly used population evidence (PM2), but newly available population evidence might come in the form of population enrichment of disease at the variant level, which is a different form of evidence (PS4). As additional submission text data becomes available, future models may be developed that can differentiate among these evidence types. Finally, the classification portion of our external validation uses threshold-derived labels from quantitative scores to represent an approximate ground truth (e.g., functional scores above a certain level represent loss of function). Although these thresholds are based on prior work, they do not capture the full nuance of expert-curated variant classifications, which may differ across genes and phenotypes. Our scoring framework should therefore be interpreted as a prioritization tool to support expert review, rather than as an automated reclassification system.

## Conclusion

In this study, we investigated the potential of using language models to extract evidence according to the ACMG evidence guidelines from text summaries on ClinVar. We used LLMs to annotate high-quality variant summaries and built the first ACMG evidence dataset, VETA. With this data set, we fine-tuned language models to classify the evidence presented in variant text descriptions.

This work addresses a bottleneck in variant interpretation: systematically integrating diverse types of evidence as it emerges. By transforming unstructured evidence summary texts into structured evidence types, our method enables scalable identification of evidence gaps, systematic integration of emerging data, and prioritization of variants for reclassification. Although automated approaches cannot replace expert clinical judgment, they can improve the efficiency and completeness of variant curation efforts.

## Data Availability

Submission-level text summaries and predictions for each evidence type are available at https://doi.org/10.6084/m9.figshare.30865748.v1 for all available ClinVar submissions with a submission summary text. A website interface for lookup prediction results is available at https://huggingface.co/spaces/weijiang99/ acmg-evidence-prediction, and prediction results can be queried via API on Hugging Face.

https://figshare.com/articles/dataset/ClinVar-BERT_Evidence_Extraction_Models_Prediction_Results_on_ClinVar_Data/30865748/1

## Author Contributions

W.L. and C.C. wrote the main manuscript text with the advice of M.L. and M.Z. V.B and T.Y provided supportive data and methods. W.L. prepared all training datasets, conducted all main analyses, and produced all figures and tables. All authors reviewed the manuscript.

## Acknowledgements

We are indebted to the UKB and its participants (UKB application 41250 and IRB protocol 2020P002093).

## Supplementary Material

### Data Annotation

We used the following prompt for annotating variant summaries using LLMs.

#### LLM Annotation Prompt

##### System Message

You are an advanced AI trained to identify and extract ACMG (American College of Medical Genetics and Genomics) evidence types from clinical genetic data summaries. Your task is to analyze a submission summary and determine whether there is any evidence or description associated with one or more ACMG evidence types. If you find any, please provide the ACMG evidence type along with its corresponding descriptive text as mentioned in the summary.

##### Content

You are an advanced AI trained to identify and extract ACMG codes from clinical genetic data comments. Your task is to analyze a submission comment and pinpoint any ACMG codes, extracting them along with their descriptive context.

##### Context

ACMG codes are used to classify genetic variants by their potential clinical impact. They follow specific formats like PS2, PS3, PM2, PP3, etc. There are 27 distinct types, including: BA1, BS1, BS2, BS3, BS4, PS1, PS2, PS3, PS4, PM1, PM2, PM3, PM4, PM5, PM6, PP1, PP2, PP3, PP4, PP5, PVS1, BP1, BP2, BP4, BP5, BP6, BP7.

##### Your Task

- Identify any ACMG codes in the submission comment, including modified forms like PM2 Supporting, PP4 Moderate, etc.
- Only extract text that is clearly labeled with an ACMG code—ignore non-ACMG labels.
- For each code, extract the exact sentence(s) or clause(s) from the submission comment that directly provide evidence for or explain the code. These descriptions should be complete, literal excerpts from the comment.
- Format your output like this: [ACMG Code]: [Exact descriptive text from the comment]
- Make sure the ACMG code is written exactly as shown in the original comment (e.g., PP3, PM2 Supporting).
- If no ACMG codes are present, respond with: “No ACMG code found.”

##### Submission comment

`{comment}`

### VETA Dataset Annotation and Distribution Statistics

**Table 1:** Agreement rate between Llama, Mistral, and GPT-4o-mini’s annotation result on ClinVar and ClinGen variant summary.

|  | GPT-4o-mini annotation on<br>ClinVar submission summary | GPT-4o-mini annotation on<br>ClinGen variant summary |
| --- | --- | --- |
| Mistral-7b | 91.94% | 92.59% |
| Llama-3.1-8b | 65.28% | 76.88% |

**Table 2:** Distribution of ACMG evidence types in the annotated data from ClinVar and ClinGen.

| ACMG Code | Count | ACMG Code | Count | ACMG Code | Count |
| --- | --- | --- | --- | --- | --- |
| PM2 | 6,465 | PVS1 | 4,585 | PP3 | 4,135 |
| PM3 | 4,110 | PS4 | 3,822 | PS3 | 3,507 |
| PP4 | 3,373 | PP1 | 2,312 | BP4 | 2,042 |
| PM1 | 1,534 | PM5 | 1,284 | BS1 | 1,247 |
| PS2 | 847 | BS2 | 786 | BA1 | 780 |
| BP7 | 746 | PP2 | 660 | PM6 | 606 |
| BS3 | 392 | PM4 | 309 | PS1 | 300 |
| BP5 | 247 | BP2 | 159 | PP5 | 97 |
| BP1 | 91 | BS4 | 63 | BP6 | 23 |

### Model Training Performance

#### VETA as Test Data

**Table 3:** Evidence classification model training results.

| Evidence | Stage | Accuracy | Precision | Recall | F1 |
| --- | --- | --- | --- | --- | --- |
| Computational | Stage 1 | 0.9842 | 0.9841 | 0.9842 | 0.9841 |
| Computational | Stage 2 | 0.9891 | 0.9891 | 0.9891 | 0.9891 |
| Functional | Stage 1 | 0.9806 | 0.9806 | 0.9806 | 0.9806 |
| Functional | Stage 2 | 0.9612 | 0.9614 | 0.9612 | 0.9612 |
| Population | Stage 1 | 0.9702 | 0.9701 | 0.9702 | 0.9701 |
| Population | Stage 2 | 0.9857 | 0.9857 | 0.9857 | 0.9857 |

#### ClinVar as Training Data, ClinGen as Test Data

**Table 4:** Evidence classification model training results. Each evidence type shows performance at Stage 1 and Stage 2.

| Evidence Type | Stage | Accuracy | Precision | Recall | F1 |
| --- | --- | --- | --- | --- | --- |
| Computational | Stage 1 | 0.9923 | 0.9776 | 0.9819 | 0.9797 |
|  | Stage 2 | 0.9968 | 0.9953 | 0.9986 | 0.9969 |
| Functional | Stage 1 | 0.9907 | 0.9034 | 0.9828 | 0.9415 |
|  | Stage 2 | 0.9662 | 0.9970 | 0.9633 | 0.9798 |
| Population | Stage 1 | 0.9656 | 0.9642 | 0.9239 | 0.9436 |
|  | Stage 2 | 0.9897 | 0.9902 | 0.9954 | 0.9928 |

### Model Evaluation

#### Stage 1 Model Evaluation Results

**Table 5:** Stage 1 Model Evaluation with ClinGen Summary Results.

| Evidence Type | Accuracy | Precision | Recall | F1 Score | AUC |
| --- | --- | --- | --- | --- | --- |
| Computational | 0.8656 | 0.8276 | 0.9274 | 0.8747 | 0.8979 |
| Functional | 0.9494 | 0.8025 | 0.8712 | 0.8354 | 0.9521 |
| Population | 0.8911 | 0.9324 | 0.9476 | 0.9399 | 0.6060 |
| Population (regex-based) | 0.9337 | 0.9792 | 0.9498 | 0.9643 | 0.8152 |

#### Stage 2 Model Evaluation Results

**Table 6:** Stage 2 Model Evaluation with ClinGen Summary Results.

| Evidence Type | Accuracy | Precision | Recall | F1 Score | AUC |
| --- | --- | --- | --- | --- | --- |
| Computational | 0.9314 | 0.9981 | 0.8802 | 0.9355 | 0.8908 |
| Functional | 0.8617 | 0.9968 | 0.8268 | 0.9039 | 0.8631 |
| Population | 0.9441 | 0.9976 | 0.9309 | 0.9631 | 0.9330 |

### Model Validation with External Data

**Table 7:** Stage 2 Model Evaluation Results.

| Evidence Type | Accuracy | Precision | Recall | F1 Score | AUC |
| --- | --- | --- | --- | --- | --- |
| Computational | 0.6152 | 0.8253 | 0.6152 | 0.6383 | 0.8677 |
| Functional | 0.7458 | 0.9351 | 0.7458 | 0.8024 | 0.8673 |
| Population | 0.7243 | 0.7651 | 0.7243 | 0.7307 | 0.7700 |

### Model Validation

**Fig. 1:**
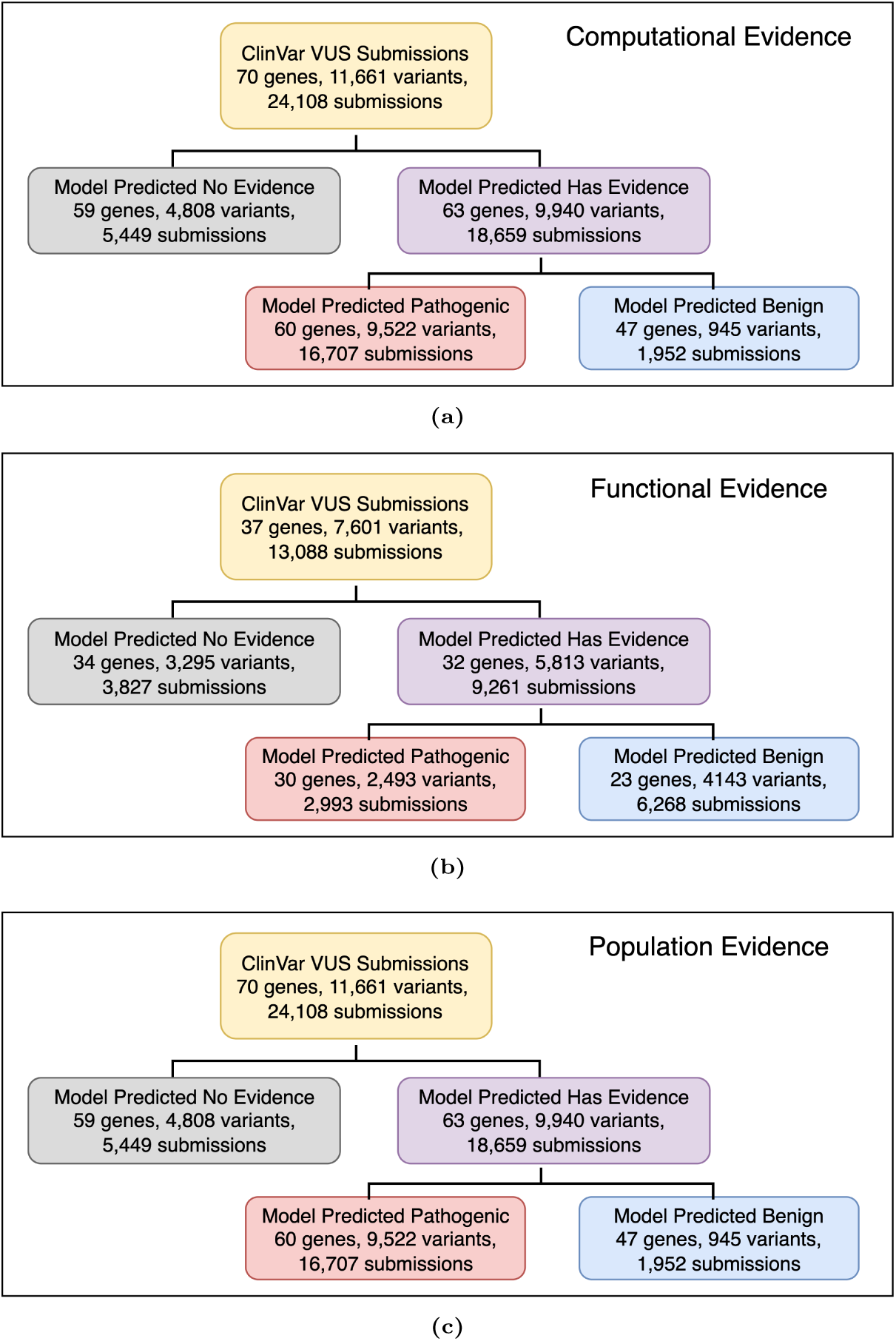
A two-stage evidence classification framework is applied across evidence types. (**a**) Computational evidence model across 70 genes (11,661 variants; 24,108 submissions). (**b**) Functional evidence model across 37 genes (7,601 variants; 13,088 submissions). (**c**) Population evidence model across 70 genes (7,601 variants; 13,088 submissions). For each evidence type, Stage 1 predicts presence/absence of evidence in the summary; Stage 2 assigns direction (pathogenic vs benign) for evidence-positive submissions.

### Additional Validation Results

**Fig. 2:**
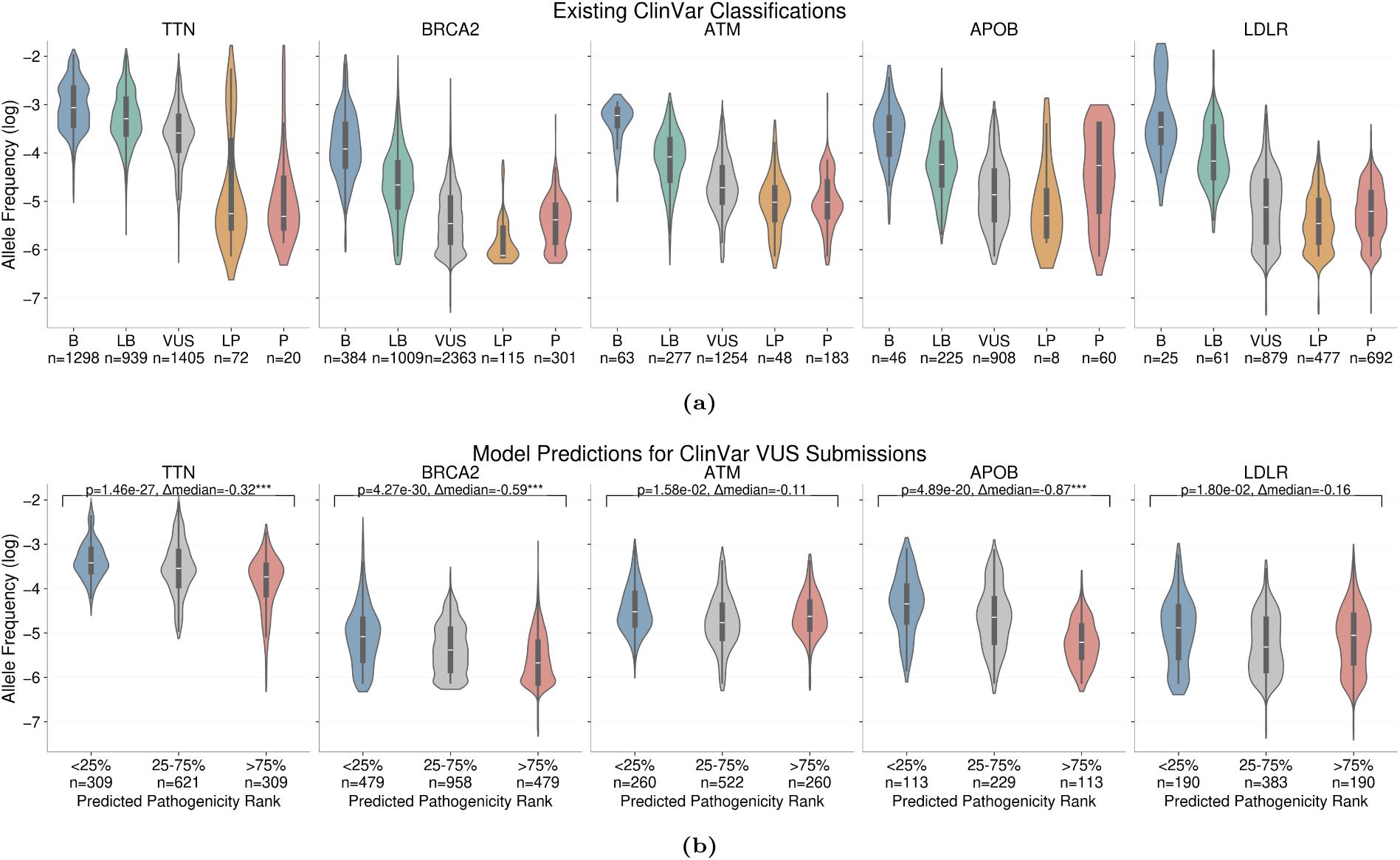
Population evidence validation of model predictions using allele frequencies. (**a**) gnomAD allele frequencies across ClinVar classifications. (**b**) gnomAD allele frequencies across model-predicted VUS categories. VUS submissions were stratified into three percentile bins, ranked by predicted pathogenicity: low (bottom 25%), intermediate (middle 50%), and high (top 25%). Statistical significance reported after Bonferroni correction (* *p<*0.05, ** *p<*0.01, *** *p<*0.001).

**Fig. 3:**
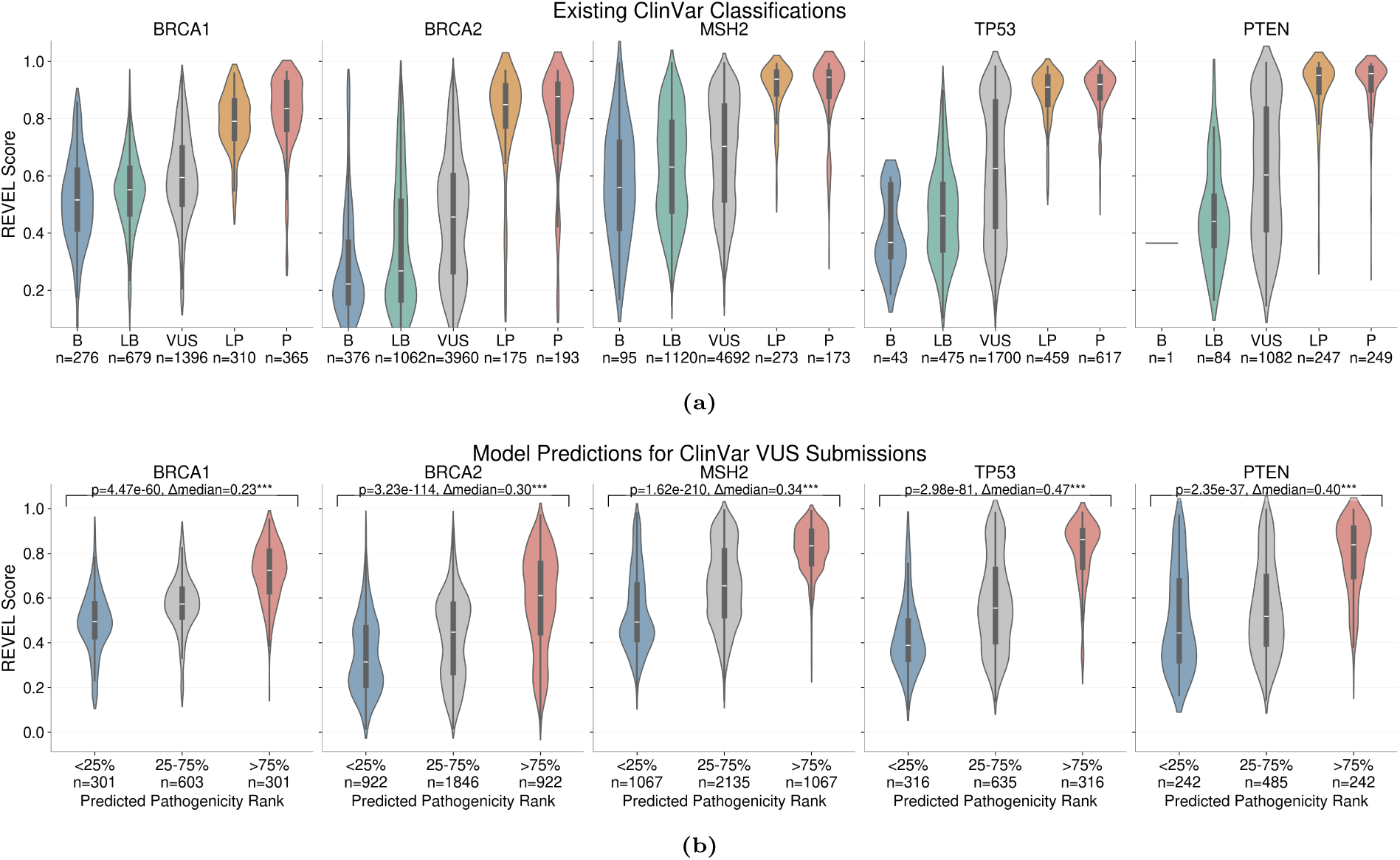
Computational evidence classification and model performance analysis. (**a**) Gene-specific violin plots of existing ClinVar submission classifications as B, LB, VUS, LP, and P on the *x*-axis and each submission’s corresponding REVEL scores on the *y*-axis. (**b**) Stage 2 model predictions for ClinVar VUS submissions stratified by confidence using percentile-based thresholds. VUS submissions were stratified into three percentile bins, ranked by predicted pathogenicity: low (bottom 25%), intermediate (middle 50%), and high (top 25%). Statistical significance reported after Bonferroni correction (* *p<*0.05, ** *p<*0.01, *** *p<*0.001).

**Fig. 4:**
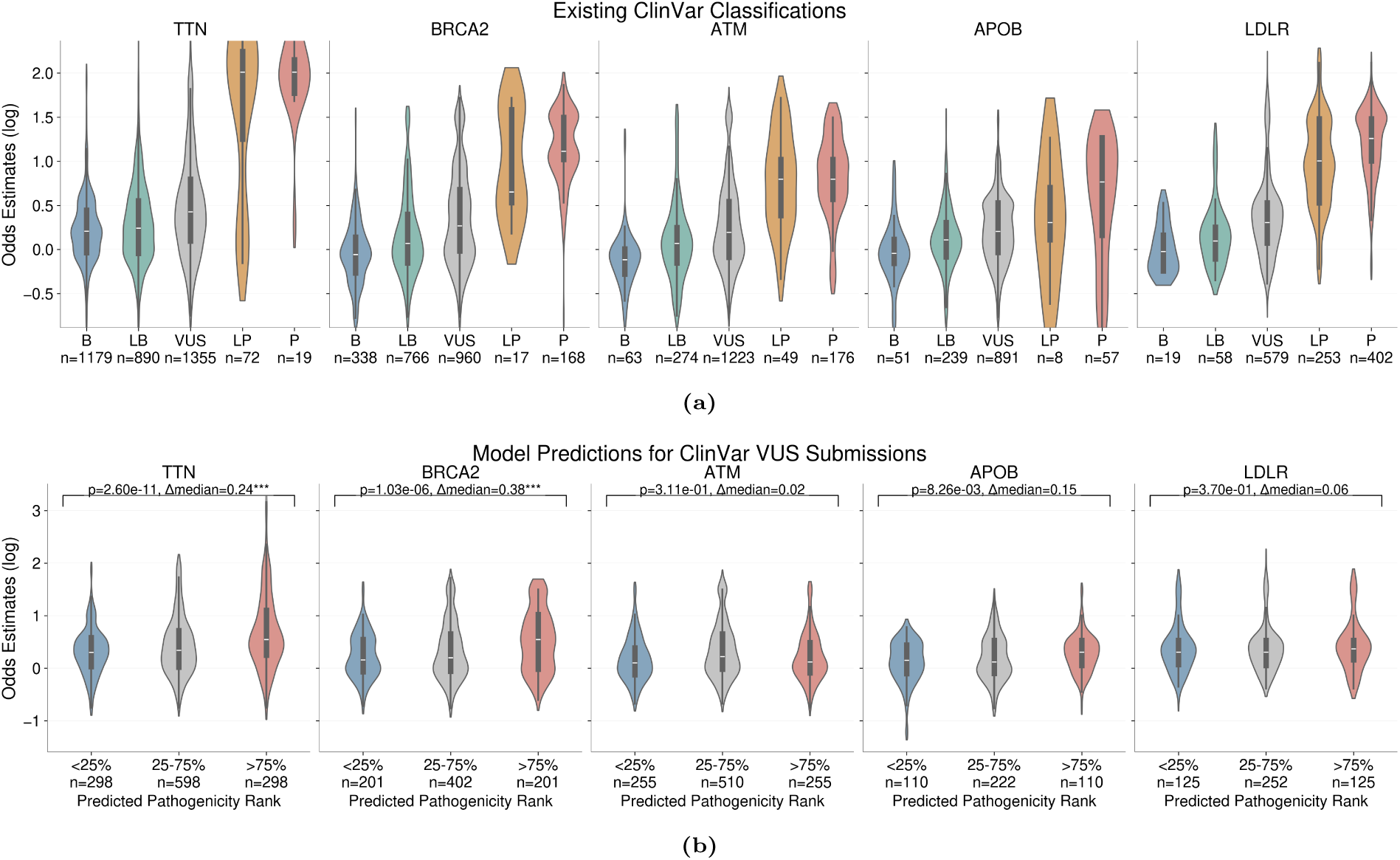
Population evidence classification and model performance analysis. (**a**) Gene-specific violin plots of existing ClinVar submission classifications as B, LB, VUS, LP, and P on the *x*-axis and each submission’s corresponding odds estimates on the *y*-axis. (**b**) Stage 2 model predictions for ClinVar VUS submissions stratified by confidence using percentile-based thresholds. VUS submissions were stratified into three percentile bins, ranked by predicted pathogenicity: low (bottom 25%), intermediate (middle 50%), and high (top 25%). Statistical significance reported after Bonferroni correction (* *p<*0.05, ** *p<*0.01, *** *p<*0.001).

#### Results for Alternative Thresholds

**Fig. 5:**
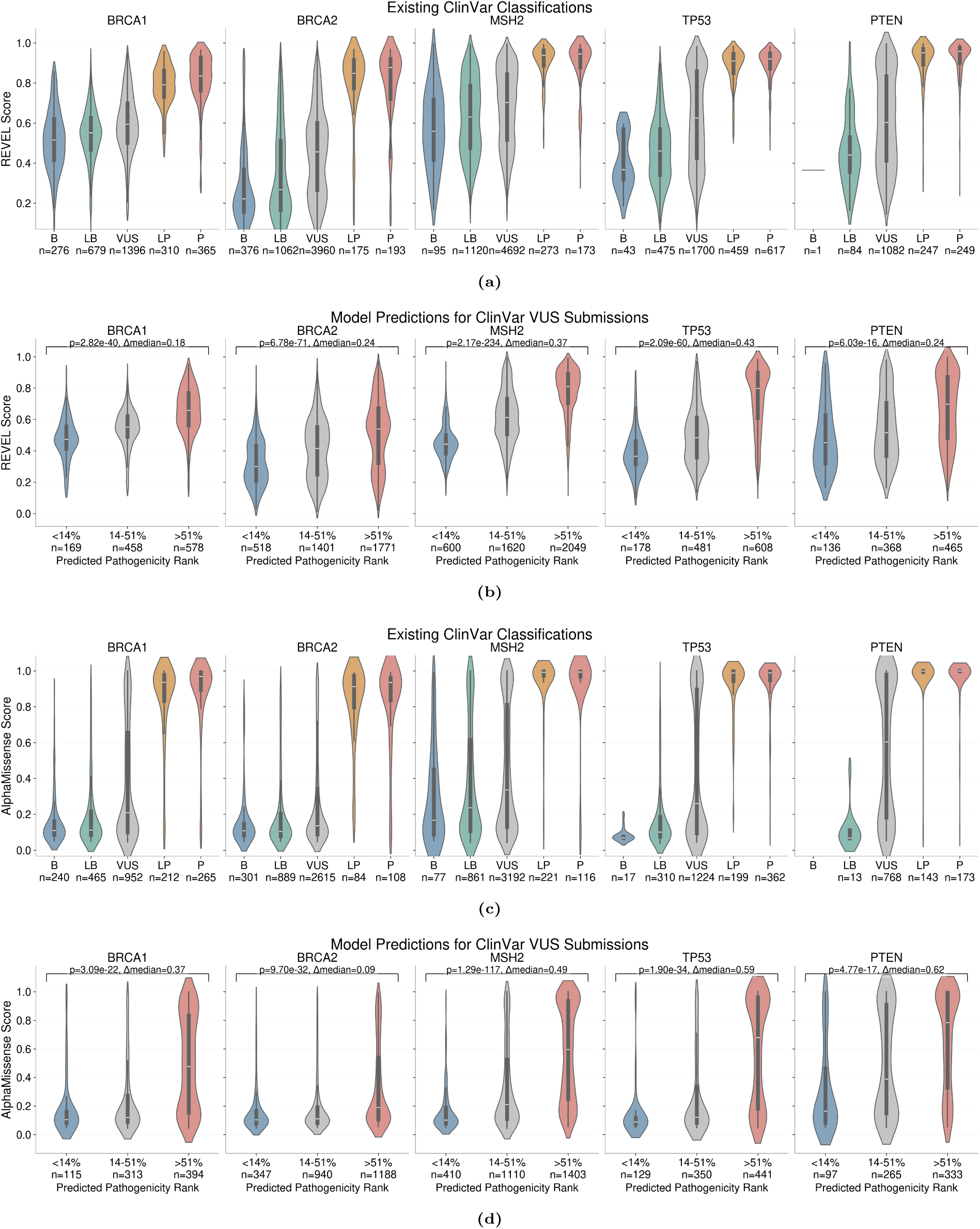
Computational evidence classification and model performance analysis. (**a**) Gene-specific violin plots of existing ClinVar submission classifications as B, LB, VUS, LP, and P on the *x*-axis and each submission’s corresponding REVEL scores on the *y*-axis. (**b**) Stage 2 model predictions for ClinVar VUS submissions stratified by confidence using percentile-based thresholds. VUS submissions were stratified by predicted pathogenicity rank using confidence-based thresholds: low (14.06%), intermediate (37.93%), and high (48.01%). (**c**) Gene-specific violin plots of existing ClinVar submission classifications as B, LB, VUS, LP, and P on the *x*-axis and each submission’s corresponding AlphaMissense scores on the *y*-axis. (**d**) Stage 2 model predictions for ClinVar VUS submissions stratified by confidence using percentile-based thresholds. VUS submissions were stratified by predicted pathogenicity rank using confidence-based thresholds: low (14.06%), intermediate (37.93%), and high (48.01%).

**Fig. 6:**
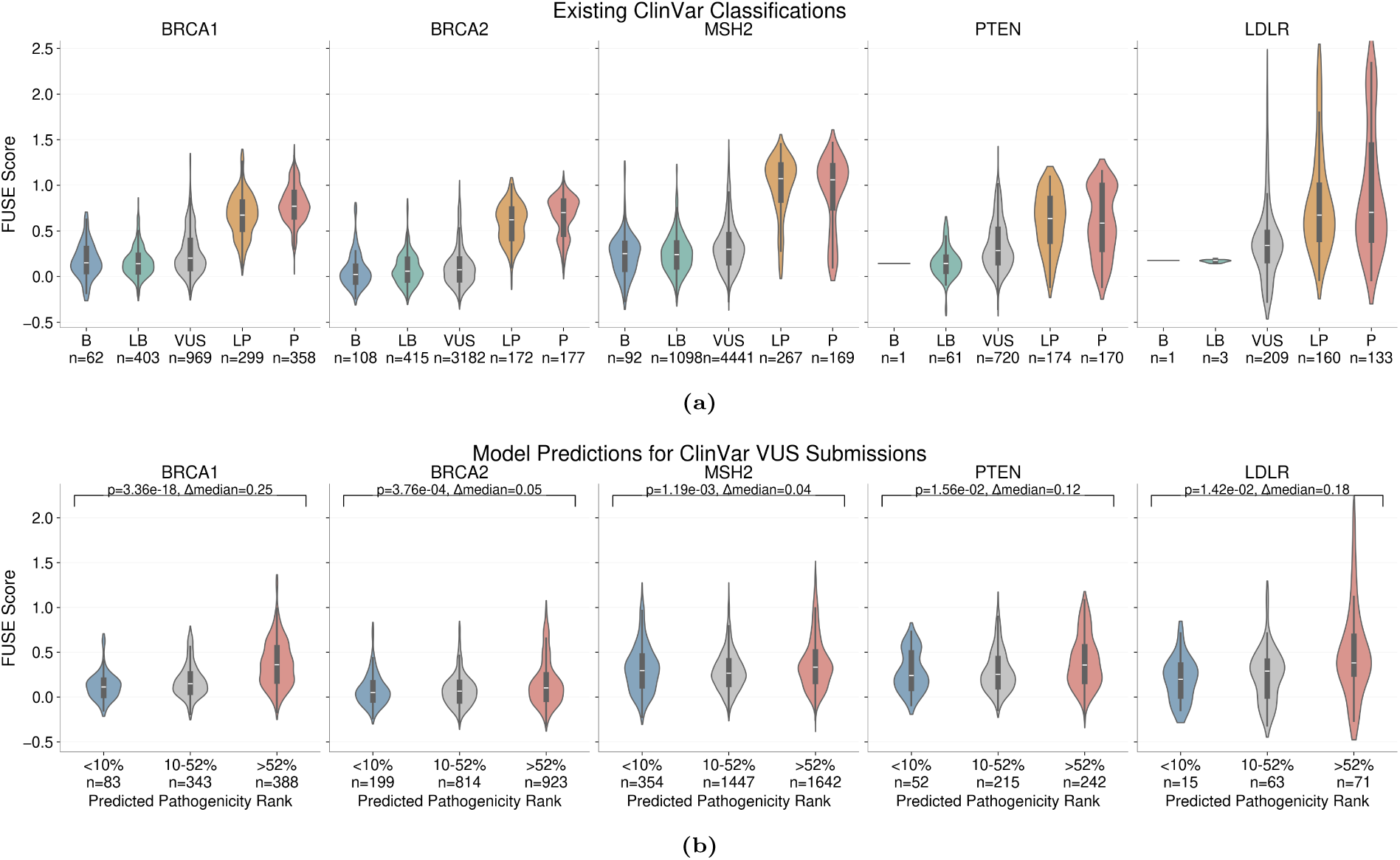
Functional evidence classification and model performance analysis. (**a**) Gene-specific violin plots of existing ClinVar submission classifications as B, LB, VUS, LP, and P on the *x*-axis and each submission’s corresponding FUSE scores on the *y*-axis. (**b**) Stage 2 model predictions for ClinVar VUS submissions stratified by confidence using percentile-based thresholds. VUS submissions were stratified by predicted pathogenicity rank using confidence-based thresholds: low (10.29%), intermediate (42.00%), and high (47.71%).

**Fig. 7:**
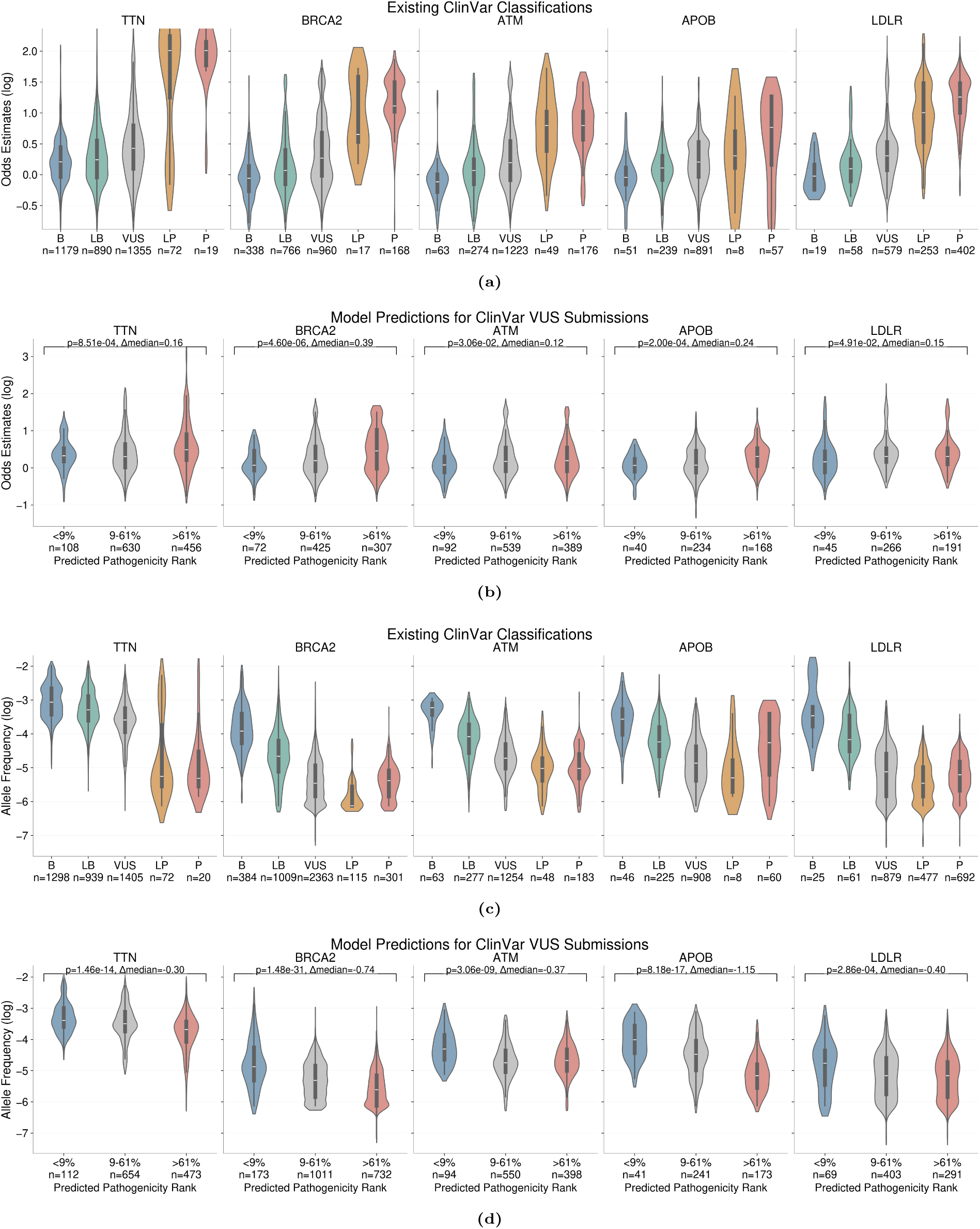
Population evidence classification and model performance analysis. (**a**) Gene-specific violin plots of existing ClinVar submission classifications as B, LB, VUS, LP, and P on the *x*-axis and each submission’s corresponding odds estimates on the *y*-axis. (**b**) Stage 2 model predictions for ClinVar VUS submissions stratified by confidence using percentile-based thresholds. VUS submissions were stratified by predicted pathogenicity rank using confidence-based thresholds: low (9.06%), intermediate (52.70%), and high (38.23%). (**c**) Gene-specific violin plots of existing ClinVar submission classifications as B, LB, VUS, LP, and P on the *x*-axis and each submission’s corresponding allele frequencies on the *y*-axis. (**d**) Stage 2 model predictions for ClinVar VUS submissions stratified by confidence using percentile-based thresholds. VUS submissions were stratified by predicted pathogenicity rank using confidence-based thresholds: low (9.06%), intermediate (52.70%), and high (38.23%).

### Model Validation with LLM

We used the following prompt for checking model predictions with LLMs.

#### LLM Validation Prompt

##### System Prompt

You are an expert in clinical genetics, specifically trained in applying ACMG guidelines for variant interpretation. Your task is to assess whether a given ClinVar variant summary contains specific types of evidence according to ACMG guidelines.

##### Content

We are trying to identify whether there are specific forms of evidence about genetic variants from clinical genetics diagnostic reports. The goal is to identify whether forms of evidence from the ACMG/AMP Sequence Variant Interpretation guidelines are present or absent in each report.

Here is a report we would like to analyze:

`{clinvar_summary}`

Does this variant summary contain variant-specific information related to the `{evidence_type}` evidence type, based on the ACMG sequence variant interpretation guidelines?

Do not assess whether the evidence supports or contradicts pathogenicity—only check for the presence of specific variant-level evidence that is directly related to the evidence type `{evidence_type}`.

The specific types of evidence and their general descriptions from the ACMG guidelines for `{evidence_type}` include:

`{format_evidence_type(evidence_type)}`

Please answer only based on whether the summary explicitly contains specific information that is analyzed in the context of this variant related to `{evidence_type}` evidence.

- If there is any variant-specific information directly related to `{evidence_type}` evidence, respond “Yes”.
- If there is no mention of information directly related to `{evidence_type}` evidence at all, respond “No”.
- Also respond “No” if there is only general information about all types of evidence reviewed, but nothing specific about this evidence type for the variant.

Provide your response in the following format:

- **Assessment:** [Yes or No]
- **Confidence:** [Score between 1–100, where 100 indicates absolute certainty]
- **Reasoning:** [Brief explanation based only on the explicit presence of `{evidence_type}` evidence, without evaluating its impact]

**Example response 1:**

`Assessment: Yes`

`Confidence: 95`

`Reasoning: The summary explicitly mentions functional studies showing loss of protein function for this specific variant, which is related to functional evidence.`

Example response 2:

`Assessment: No`

`Confidence: 90`

`Reasoning: The summary does not mention any information related to allele frequency or population enrichment of cases, which is related to population evidence.`

**Fig. 8:**
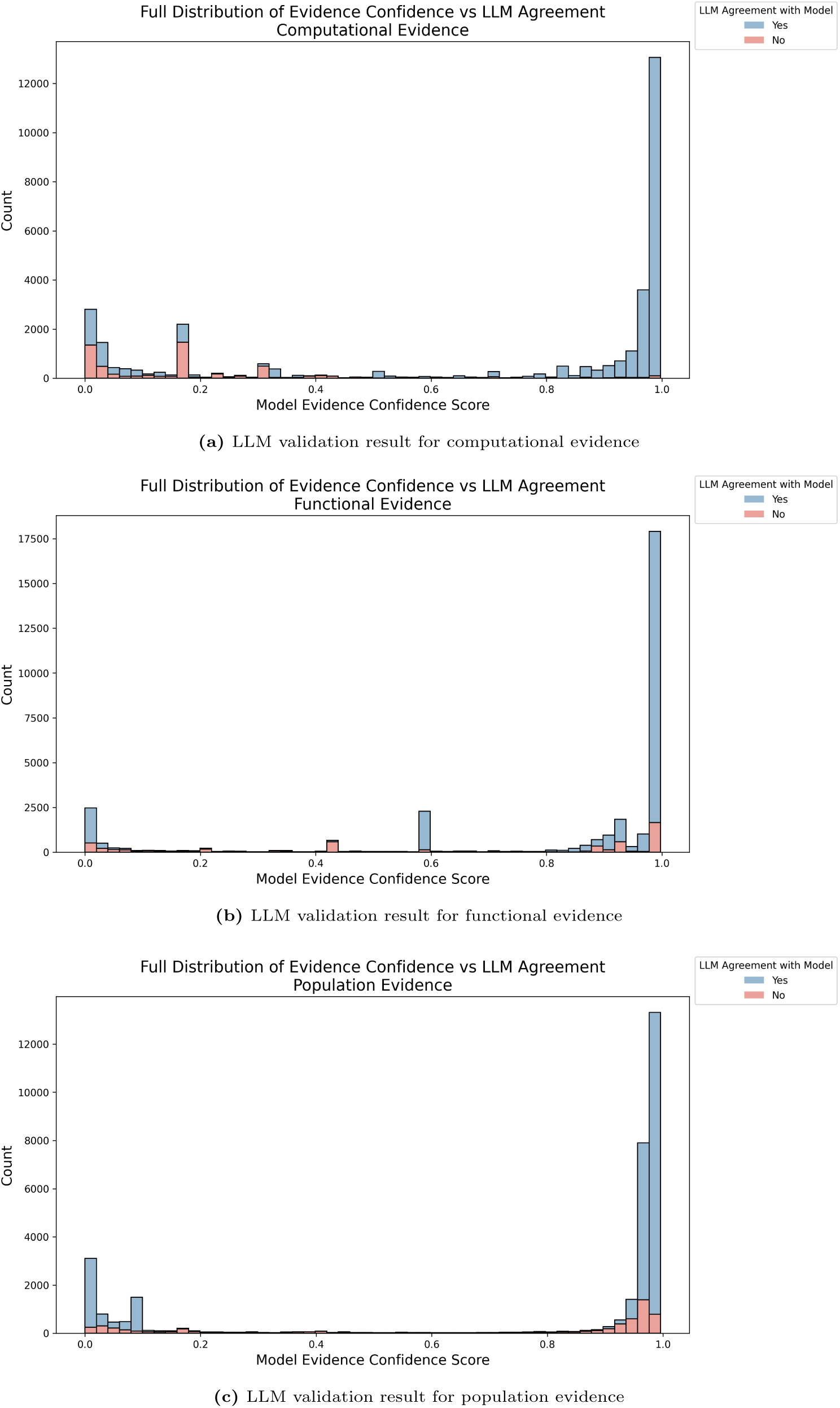
Results for LLM validation using Llama-3.1-8b as the judge to validate model prediction results. Here we show LLM validation results for all the prediction results from the computational, functional, and population classifiers. We made the histogram for all model prediction confidence scores, with the blue-colored bar representing LLM responses that agree with the model prediction, and the red-colored portion representing LLM responses that disagree with the model prediction. (**a**) LLM validation result for computational evidence classifier predictions results; (**b**) LLM validation result for functional evidence classifier predictions results; (**c**) LLM validation result for population evidence classifier predictions results;

### Identifying Variants with Evidence Gaps

#### Variant annotations

The canonical functional consequence of each variant was calculated using Variant Effect Predictor (v108) [28]. Non-coding variants outside of essential splice sites were not considered in the analysis. Non-synonymous coding variants were included with any of the following canonical consequences:

- splice acceptor variant
- splice donor variant
- stop gained
- frameshift variant
- stop lost
- start lost
- missense variant
- inframe insertion
- inframe deletion

REVEL scores were aggregated from all available transcripts and annotated if available in at least one transcript.

**Table 8:** Odds ratio thresholds for pathogenic PS4 evidence applied from *Bhat, et al* [. **11]**

| Phenotype | Supporting (+1) | Moderate (+2) | Strong (+4) | Very Strong (+8) |
| --- | --- | --- | --- | --- |
| Familial hypercholesterolemia | 5.2–8.1 | 8.1–49.6 | $\geq 49.6$ | |
| Breast and ovarian cancer | 4.2–5.4 | 5.4–13.7 | $\geq 13.7$ | |
| Colorectal cancer | 8.5–12.0 | 12.0–17.6 | 17.6–20.0 | $\geq 20.0$ |
| Maturity-onset diabetes young | 17.6–18.7 | 18.7–21.8 | 21.8–24.6 | $\geq 24.6$ |
| Primary familial hypertrophic cardiomyopathy | 73.6–74.4 | 74.4–84.0 | 84.0–102.6 | $\geq 102.6$ |
| Primary dilated cardiomyopathy | 29.6–57.9 | $\geq 57.0$ | | |
| Arrhythmogenic right ventricular cardiomyopathy | 14.6–18.6 | 18.6–60.5 | 60.5–131.3 | $\geq 131.3$ |
| Li-Fraumeni syndrome | 2.6–3.4 | 3.4–6.2 | $\geq 6.2$ | |
| Default (other phenotypes) | 6.7–12.0 | 12.0–40.9 | $\geq 40.9$ | |

**Table 9:** Allele frequency thresholds for benign evidence.

| Phenotype | Strong benign (-4) | Very strong benign (-8) |
| --- | --- | --- |
| Arrhythmogenic right ventricular cardiomyopathy | $> 1e-3$ | $> 5e-2$ |
| Primary dilated cardiomyopathy | $> 4e-3$ | $> 5e-2$ |
| Breast and ovarian cancer | $> 1.25e-1$ | $> 5e-2$ |
| Colorectal cancer | $> 4.3e-2$ | $> 5e-2$ |
| Familial hypercholesterolemia | $> 4e-3$ | $> 5e-2$ |
| Primary familial hypertrophic cardiomyopathy | $> 2e-3$ | $> 5e-2$ |
| Brugada syndrome | $> 5e-4$ | $> 5e-2$ |
| Ehlers-Danlos syndrome type 4 | $> 2e-5$ | $> 5e-2$ |
| Juvenile polyposis syndrome | $> 1e-5$ | $> 5e-2$ |
| Romano-Ward syndrome | $> 5e-4$ | $> 5e-2$ |
| Long QT syndrome | $> 4e-4$ | $> 5e-2$ |
| Maturity-onset diabetes young | $> 1e-4$ | $> 5e-2$ |
| Osler hemorrhagic telangiectasia syndrome | $> 2e-4$ | $> 5e-2$ |
| Retinitis pigmentosa | $> 2.5e-4$ | $> 5e-2$ |
| Drash syndrome | $> 1e-6$ | $> 5e-2$ |
| Carcinoma of the pancreas | $> 1.6e-2$ | $> 5e-2$ |
| Hereditary hemochromatosis | $> 5e-3$ | $> 5e-2$ |
| Muir-Torre syndrome | $> 3.6e-3$ | $> 5e-2$ |
| Li-Fraumeni syndrome | $> 2e-4$ | $> 5e-2$ |
| Malignant tumor of the prostate | $> 1.25e-1$ | $> 5e-2$ |
| Default (other phenotypes) | $> 1e-5$ | $> 5e-2$ |

**Table 10:** REVEL score thresholds applied from *Pejaver et al* [. **23]**

| Category | Threshold | Points |
| --- | --- | --- |
| Strong pathogenic | $\geq 0.932$ | +4 |
| Moderate pathogenic | 0.773–0.931 | +2 |
| Supporting pathogenic | 0.644–0.772 | +1 |
| Supporting benign | 0.290–0.183 | -1 |
| Moderate benign | 0.183–0.016 | -2 |
| Strong benign | 0.016–0.003 | -4 |
| Very strong benign | $\leq 0.003$ | -8 |

## Additional Results

**Fig. 9:**
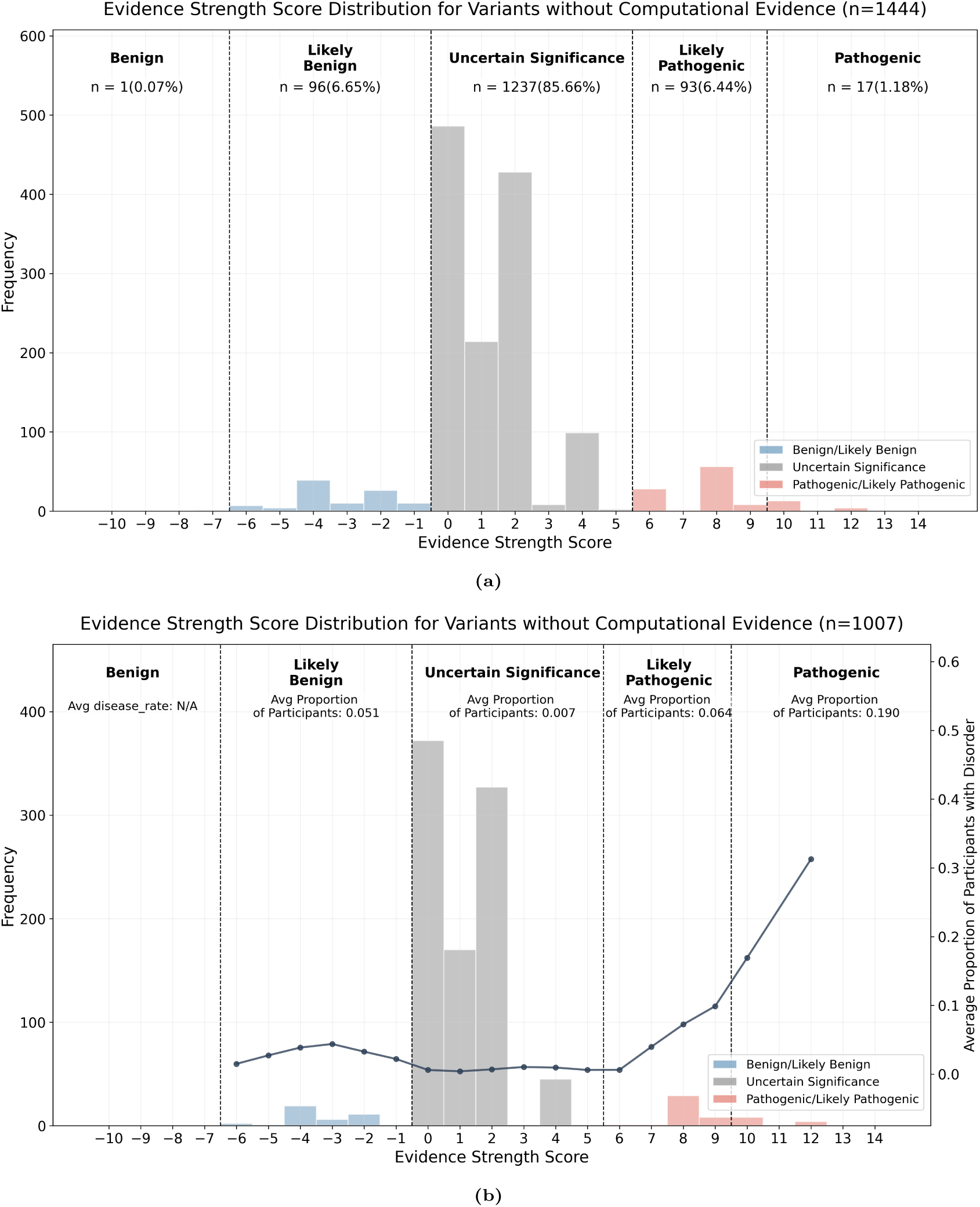
(**a**) Overall distribution of evidence strength scores across all 1,444 variants classified to have no computational evidence by the computational evidence classifier. (**b**) Distribution of evidence strength scores across 1,007 variants classified to have no computational evidence by the computational evidence classifier with phenotype and patient data available.

## Footnotes

1 https://erepo.clinicalgenome.org/evrepo/

